# Implementation of Shared Decision-Making in the Management of Chronic Musculoskeletal Pain: *a scoping review*

**DOI:** 10.1101/2025.10.02.25336876

**Authors:** Liv Nyhave Andersen, Alex Waddell, Laura Boland, Michael Skovdal Rathleff, Malene Plejdrup Hansen, Janus Nikolaj Laust Thomsen, Glyn Elwyn, Jette Frost Jepsen, Kristian Damgaard Lyng

## Abstract

Shared decision-making (SDM) is increasingly recommended for managing chronic musculoskeletal pain, yet its use and implementation in clinical practice remains poorly understood. This scoping review aimed to identify and synthesize barriers and facilitators to the implementation and use of SDM across healthcare settings. A systematic search of seven databases conducted in April 2025 yielded 28 eligible studies. Using a deductive–inductive–deductive analysis approach, we mapped findings to existing SDM taxonomies and the Theoretical Domains Framework. We identified 16 themes and 46 subthemes spanning patient-, clinician-, interactional-, and system-level factors. Key facilitators included SDM training, decision aids, effective communication, empathetic care, trust, and strong therapeutic alliances. Barriers included time constraints, lack of individualized care, insufficient knowledge, conflicting beliefs about pain, and system-level obstacles such as limited resources or organizational support. Prominent theoretical domains included knowledge, skills, beliefs about consequences, and environmental context and resources. These findings offer a comprehensive overview of multilevel factors shaping SDM in musculoskeletal care. Future studies should focus on developing context-sensitive knowledge translation interventions to overcome barriers and leverage facilitators to promote the uptake of SDM in musculoskeletal pain.

**Trial registration:** https://doi.org/10.17605/OSF.IO/SH8G4.

## 1. Introduction

Chronic musculoskeletal pain is one of the main global contributors to disability and poor health status [9]. Chronic musculoskeletal pain affects one in every three people and is associated with anxiety, depression, insomnia, and fatigue [17,35]. The combined direct and indirect costs of chronic musculoskeletal pain amass up to 2% gross national product in Europe [6]. Management of chronic musculoskeletal pain is a complex task, requiring evidence-based treatment tailored to the individual’s needs [25]. However, no single treatment or combined treatment program has demonstrated clear superior effectiveness, and most of the research in this domain is of low quality [57]. In the face of uncertainty about the most effective treatment, shared decision-making (SDM) offers a valuable framework for supporting patients’ informed decision [26,27]. SDM involves collaborative discussions between patients and healthcare professionals, focusing on the effectiveness, risks, and benefits of available options while aligning these with patient preferences [28]. Evidence suggests that SDM can enhance patient knowledge, improve risk perception, foster active involvement, and reduce decisional conflict [34,38,74,83]. Despite its demonstrated benefits, the implementation and use of SDM in the management of chronic musculoskeletal pain remains limited [37,70,79]. Research from health domains has identified the barriers and facilitators to the implementation of SDM in clinical practice [2,10,50,65,77,84]. However, the successful implementation of SDM might also be influenced by broader systemic factors, including healthcare policies, institutional support, and cultural attitudes towards patient autonomy and involvement in decision-making [37,46]. Addressing these specific barriers and facilitators has been shown to enhance SDM compared to usual care across various settings [49]. Therefore, this scoping review aims to describe the barriers and facilitators of SDM and its implementation in the management of chronic musculoskeletal pain across healthcare sectors.

## 2. Methods

### 2.1. Protocol and Registration

This scoping review followed the framework proposed by Arksey and O’Malley [4], refined by Levac et al. [53], and guided by the Joanna Briggs Institute methodology for scoping reviews [66]. The reporting of this scoping review followed the Preferred Reporting Items for Systematic review and Meta-Analysis (PRISMA-ScR) extension for scoping reviews [80]. The protocol was pre-registered on Open Science Framework (DOI: 10.17605/OSF.IO/QM5WN, [3]).

### 2.2. Protocol Deviation

This study included one deviation from the original protocol in which our study was converted from a systematic review to a scoping review. The scoping review approach was deemed more fitting to answer the aim(s) of this study, and the limited available evidence [60]. This deviation did not affect the approach to data analysis but led to a decision to leave out critical appraisal of the included studies since this scoping review describes the available evidence, identifies key characteristics or factors related to a concept (i.e., SDM), and analyses knowledge gaps, as suggested by Munn et al [60].

### 2.3. Information Sources and Search

The literature search was designed by an experienced research librarian (JFJ) with inputs from the entire research team. The search string was adapted from similar reviews with different settings of barriers and facilitators within the implementation of SDM [45,50]. The literature search was conducted in April 2025 in the following databases: PubMed, Embase (Embase.com), The Cochrane Library, CINAHL (EBSCOhost), Scopus, PEDro, and Psy-cINFO (APA). Controlled vocabulary, e.g. Medical Subject Headings (MeSH), was used in combination with text words related to “Shared Decision-Making” and “Musculoskeletal pain”. The search included all articles related to existing facilitators and barriers for chronic musculoskeletal pain-focused SDM management and the implementation of these. See example of literature search in PubMed in **Supplementary Appendix 1**.

### 2.4. Eligibility Criteria

Population, Intervention, Comparison, Outcomes and Study (PICOS) was used to define in– and exclusion criteria (see **TABLE 1)** and was used to search for both quantitative and qualitative studies [59]. Included participants were either authorized healthcare practitioners, carer/relatives, adults (aged 18 or above) diagnosed with any type of chronic musculoskeletal pain, or observers (i.e., individuals not directly involved with the SDM process but through evaluation or similar). Studies that comprised an SDM intervention were included, which also covered decision aids and similar. All study types with original data were included, meaning that both qualitative and quantitative studies were included. Data related to either barriers or facilitators of SDM were included. No limit on publication date was applied; however, the included articles were restricted to English language only.

### 2.5. Selection of Sources of Evidence

EndNote 20 (Clarivate Analytics, Philadelphia, United States) was used to remove duplicates, and Covidence software (Covidence Systematic Review Software, Veritas Health Innovation, Melbourne, Australia) was used to manage all records. Two reviewers (LNA and KDL) independently screened titles and abstracts, followed by full-text screening of all potentially eligible studies. Disagreements at any stage were first resolved through discussion between the two reviewers. If consensus could not be reached, a third author (MSR or JNLT) was available to make the final decision.

### 2.6. Data Charting

Prior to data extraction, internally iterated and purpose-built extraction forms were developed and piloted. Data extraction included study characteristics such as author, year of publication, country, title, journal, study aim, study design, method of data collection, and participants. Additionally, we extracted data on barriers and facilitators using NVivo 12 (QSR International Pty Ltd. V12, 2020). Two reviewers (LNA and KDL) independently extracted data from 20% of included studies to ensure consistency, proceeding only after reaching at least 90% agreement. The remaining studies were extracted by LNA and independently verified for accuracy and completeness by KDL or another senior author.

### 2.7. Data Analysis

Barriers and facilitators were analysed using a three-step deductive-inductive-deductive content analysis. First, we mapped the barriers and facilitators onto pre-existing taxonomies related to SDM formulated by Légaré et al. 2008 and Joseph-Williams et al. 2014, which consisted of 38 and 18 factors, respectively [45,50]. Second, to conduct the inductive coding, we vertically (i.e., across the individual papers) identified themes from all included articles using direct quotes, where possible. This was followed by a horizontal coding (i.e., across the entire data set) comparing and combining themes across articles with similar semantics. Finally, the results from the first two steps were mapped onto the Theoretical Domains Framework (TDF) [5,13]. To combine counts of barriers and facilitators from both qualitative and quantitative studies, each article was counted once using the following criteria:

- If one paper reported the same barrier or facilitator multiple times, they will be counted only once
- If one paper reported the same factor as both a barrier and a facilitator, it will be counted once as a barrier and once as a facilitator
- If one paper reported multiple perspectives (e.g., HCPs and patients), and each participant type reports the same barrier or facilitator, it will be counted once for each participant type [10].

## 3. Results

### 3.1. Selection of Sources

The literature search identified 6439 studies, of which 2380 duplicates were removed. The initial title and abstract screening resulted in the exclusion of 4059 studies, leaving 115 full-text studies to be assessed for eligibility. A total of 83 studies were excluded by full-text screening, leaving 28 studies for data extraction and analysis [1,7,8,11,12,15,18,19,21,23,24,30,31,33,39,41,42,44,47,48,52,54–56,58,63,64,69,71,72,78]. See **Figure 1** for the PRISMA flowchart.

**FIGURE 1.**
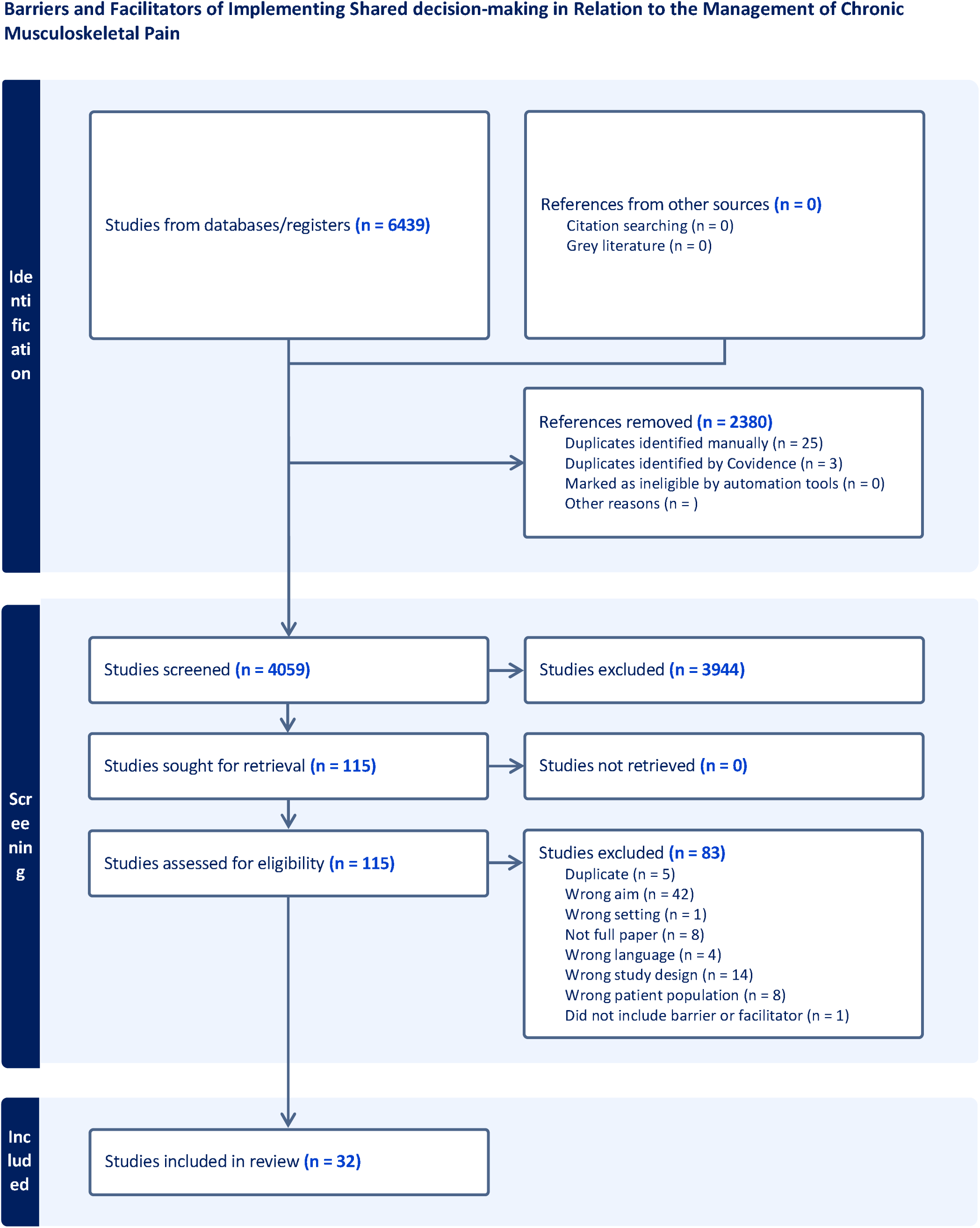
PRISMA FLOWCHART.

### 3.2. Characteristics of Sources

The included studies are described in detail in **Supplementary Appendix 2**. Fourteen different countries were represented, where USA was the most represented country (n=7), followed by England (n=6), Canada (n=4), Germany (n=3), Australia (n=2), The Netherlands (n=2), Denmark (n=1), China (n=1), Norway (n=1), Scotland (n=1), Switzerland (n=1), Taiwan (n=1), Iran (n=1) and, Saudi Arabia (n=1). Studies were published between 2006 and 2025. Of the included studies, 13 were qualitative-only study designs, primarily based on semi-structured interviews. A quantitative-only study design was used in 12 studies, with a randomized trial being the most frequent study design (n=11). In seven studies, a mixed-methods approach was used, most commonly a mixture of interviews and questionnaires. Across all studies, 16 studies focused on patients-only, five studies focused on HCPs-only, and eleven studies focused on multiple populations. In total, 3967 patients and 517 HCPs were included.

### 3.3. Coding *a priori*

Open coding onto the *a priori framework* led to references in 27/38 factors related to HCP barriers and facilitators and references in 18/18 factors related to patients’ barriers and facilitators. Apart from the factors of an undefined category for later inductive coding, the factors with the highest number of articles referenced related to HCP barriers and facilitators were “clinical situation”, “asking patients about values or clarifying values”, “organizational constraint”, “patient characteristics” and “sharing responsibility with the patient”. The highest number of articles referenced related to patient barriers and facilitators were “trust”, “expectations of outcome”, “knowledge about disease”, “condition”, “treatment options and outcomes”, “power imbalance in the patient-clinician relationship”, and “decision support”.

### 3.4. Inductive Coding

Through an iterative process of inductive coding, the data were systematically analyzed and organized into a hierarchical framework comprising 16 main themes and 49 subthemes. These themes emerged organically from the data, reflecting the breadth and complexity of the reviewed literature. The final thematic structure, including the relationships between main themes and their corresponding subthemes, is presented in **Figure 2**. Our inductive analysis are synthesized below and tabularized in **Supplementary Appendix 3**. The synthesis is grouped by *patient-level factors*, *clinician-level factors*, *interactional dynamics,* and *system-level factors*. While presented separately, many themes span multiple levels, underscoring the interrelated nature of factors influencing the use and implementation of SDM. A more granular exploration of themes and subthemes is presented in **Supplementary Appendix 4**.

**FIGURE 2.**
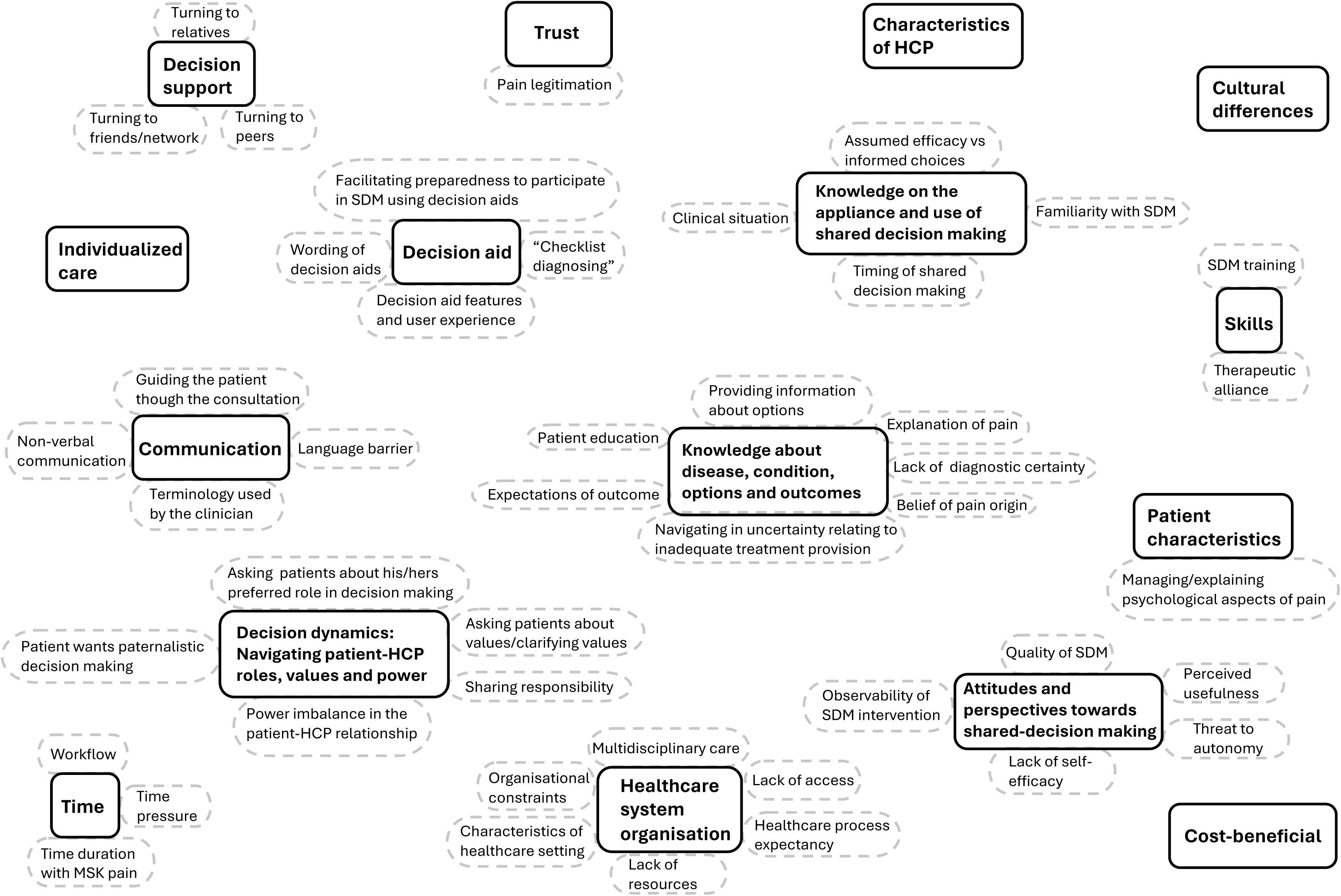
OVERVIEW OF MAIN AND SUBTHEMES.

### 3.5. Patient-level factors

#### 3.5.1. Cultural differences

This theme, represented only by patients, was reported as both a barrier and a facilitator. Facilitators included culturally competent providers who offered emotionally tailored decisions and decision aids designed to provide patient education [41,69]. Micro-cultural differences were reported as both a barrier and a facilitator to SDM, as these differences influenced trust in one’s doctor and participation in decision-making [12].

#### 3.5.2. Decision support

This theme, represented only by patients, included three subthemes: “turning to relatives,” “turning to peers,” and “turning to friends/network.” Each subtheme included both barriers and facilitators. For “turning to relatives” barriers involved pressure or conflicting beliefs[69], while facilitators included emotional reassurance and support in decisions affecting [69]. Both a barrier and facilitator were the need for relatives to reassure patients, especially when decisions could impact functional ability [69]. The subtheme “turning to peers” was reported as both a barrier and a facilitator [69]. A facilitator was when patients received guidance from others with similar pain experiences[69], but a barrier when advice was sought solely from those with similar pain [66]. The subtheme “turning to friends/network” was also both a barrier and facilitator [69]. Facilitators such as informal advice or input from medically trained friends, while formal network channels for high-stake treatments acted both as barrier and facilitator[69].

#### 3.5.3. Individualized care

This theme, represented by both patients and (HCPs), had one sub-theme: “Concern of adverse events.” Facilitators to SDM included support adjusted to different stages of treatment, management and goal setting congruent with lifestyle, personal needs, and life situation, as well as addressing issues beyond just pain [18,21,33]. Receiving a diagnosis was seen by patients as both a barrier and a relief, providing clarity for some, but for others feeling insufficient, stigmatizing, and leading to standardized rather than individualized care [11]. Patient barriers included a lack of individualized care, particularly in group rehabilitation settings [18,33,48]. It was also considered a barrier when HCPs did not address how pain affected psychosocial factors during consultations, such as daily life situations, job, family, and quality of life [48]. Both patients and HCPs expressed concerns about potential adverse events associated with different treatment options, and these concerns often differed between stakeholders depending on the type of intervention [54]. Rather than fostering open dialogue, these concerns sometimes led to avoidance of certain options, hesitancy to explore alternatives, or prematurely biased decisions, thereby constraining the scope of SDM rather than preventing it outright.

#### 3.5.4. Knowledge about disease, condition, options, and outcomes

This theme, primarily represented by patients, included six subthemes: “Patient education,” “Explanation of pain,” “Lack of diagnostic certainty,” “Belief of pain origin,” “Providing information about options,” “Expectations of outcome,” and “Navigating inadequate treatment provision.” Facilitators to SDM included educational materials and explanations in layman’s terms, which helped patients understand their condition and engage in decisions [11,18,19,21,24,48,58,78]. HCPs also considered patient education a facilitator for cooperation and engagement [58,68]. Barriers occurred when explanations were insufficient or when patients already knew too much to gain new insights [78]. “Explanation of pain” was primarily a barrier when pain causation was inconsistent or unclear, leading to frustration and differing beliefs among patients [18,48,63]. HCPs also identified this as a barrier [56]. “Lack of diagnostic certainty” was a barrier when no cause for pain was found, or when patients felt pressured to convince HCPs of their pain [48,63]. However, receiving a diagnosis could be a facilitator if HCPs asked about previous advice [63]. “Belief of pain origin” acted as both barrier and facilitator, depending on alignment between patient and HCP beliefs. Discrepancies led to delegitimization, while congruent beliefs facilitated SDM [11,21,63]. “Providing information about options” was a facilitator when HCPs provided empathetic, clear, and credible options [21,48,69]. Barriers arose when options weren’t explained, or the cause of pain was unknown [18]. “Expectations of outcome” were influenced by self-perception, age, and belief of pain origin, with mismatches in expectations being a barrier to SDM [24,48,63]. Finally, “Navigating inadequate treatment provision” was a barrier when HCPs felt pressured to provide a diagnosis or treatment despite limited options [63,78].

#### 3.5.5. Patient characteristics

This theme, represented by both HCPs and patients, included the subtheme: “Managing/explaining psychological aspects of pain”. Patient characteristics were barriers to SDM for both groups. Patients cited limited resources or financial constraints [11], while HCP noted psychological overlay, non-compliance, lack of motivation, low education, lack of acceptance of the condition, mistrust in clinicians, language or cultural barriers, and reluctance to engage in SDM [8,23]. The subtheme “managing/explaining the psychological aspects of pain” was both a barrier and a facilitator. Patients felt psychological explanations could delegitimize their pain or conflicted with biomedical views [11,48,63], but also saw value in such discussions [63,72]. Barriers also arose when, HCPs failed to address the psychosocial factors or the pain’s impact on daily life [11,63].

#### 3.5.6. Time

This theme, represented by both patients and HCPs, included three subthemes: “workflow,” “time duration with musculoskeletal pain,” and “time pressure.” The subtheme “workflow” was only represented by patients. Barriers included short appointment times [11,33,55] and long waiting times [33]. Longer consultation times were considered a facilitator [19]. The subtheme “time duration with MSK pain” was represented by both patients and HCPs. Patients saw it as both a barrier and facilitator, particularly in how age-related perceptions of health and pain acceptance influenced their engagement with SDM [63]. The duration of the pain and the length of the patient-HCP relationship were also considered both a barrier and facilitator by both groups [63]. The subtheme “time pressure” was reported by both patients and HCPs and was considered only a barrier to SDM. HCP-related barriers were influenced by limited time, with SDM seen as more time-consuming. Patients also viewed time pressure as a constraint on building a partnership with the HCP [11,23,30,33,42,68].

#### 3.5.7. Trust

This theme, reported by both patients and HCPs, included two subthemes: “stigma” and “pain legitimation.” Trust was reported by both patients and HCPs as both a barrier and facilitator. Patient barriers included losing trust in HCPs when previous treatment management was ineffective [33]. Facilitators included emotional support, taking time to discuss management, empathetic interactions, and honesty in identifying relevant treatment options within limited time [11,12,18,69]. HCP facilitators involved building rapport, while barriers arose when patients did not trust their HCPs [23,30]. The subtheme “pain legitimation” (i.e., acknowledging the patient’s experience of pain) was represented by both patients and HCPs. Both groups viewed pain legitimation as a facilitator of SDM [63]. Patient facilitators included knowledgeable, respectful, and trustworthy HCPs who acknowledged the patient’s pain experience [21,33]. Barriers included the pressure of convincing the doctor of their pain, feeling disbelieved, and being told to live with the pain [48]. HCP facilitators involved expressing empathy and validating the pain as real [48]. Both patients and HCPs agreed that pain legitimation facilitated SDM [63].

### 3.6. Clinician-level factors

#### 3.6.1. Attitude and perspectives towards shared decision-making

This theme, primarily represented by HCPs [8,11,23,24,30,42,56,63,72,78] included five subthemes: “quality of SDM,” “perceived usefulness,” “threat to autonomy,” “lack of self-efficacy,” and “observability of SDM intervention.” The subtheme “quality of SDM” was considered both a barrier [42] and a facilitator [8]. “Perceived usefulness” was primarily a facilitator, with HCPs believing that SDM would lead to improved patient outcomes and feeling optimistic about its effectiveness [23,24,56]. The intention to use SDM was both a facilitator and barrier [42], with the barrier being the belief that SDM increased service risks and was inferior to existing practices. Another barrier was disagreement with SDM principles and a preference for paternalistic care [78]. The subthemes “threat to autonomy” and “lack of self-efficacy” were barriers to SDM. HCPs reported concerns about undermining professional autonomy through SDM [30,42,78]. Lack of self-efficacy was linked to not feeling competent to perform SDM, insufficient explanations, lack of skills, and anxiety when using SDM with patients with psychological overlay [23,63,78]. Facilitators related to “observability of SDM intervention” included the ability of HCPs to observe patient benefits from SDM [23], Barriers included the lack of evaluation of SDM interventions, leading HCPs to apply it based on their own preferences [68].

#### 3.6.2. Characteristics of HCP

This theme was only reported by patients and did not generate any subthemes. Patients considered it a barrier when HCPs were abrupt during the encounter [18]. Additionally, patients considered the length of the relationship with the HCP as both a barrier and a facilitator [63]. Patient-related facilitators were related to empathetic behaviour, showing emotional support, demonstrating professional and respectful behaviour, and acknowledging and believing the patient’s pain experience [18,21,33,69].

#### 3.6.3. Communication

This theme, represented by both healthcare providers and patients, included four subthemes: “guiding the patient through the consultation,” “terminology used by clinicians,” “non-verbal communication,” and “language barriers” [18,23,30,33,48,55,69,72]. Communication was seen as both a facilitator and barrier, depending on the techniques used. Patient facilitators included well-explained treatments and clear communication [33,55]. HCP facilitators included SDM training and patient agreement with treatment options [30]. Barriers included lack of communication skills and insufficient explanations [23,55]. “Guiding the patient through the consultation” was both a facilitator and barrier. Patient facilitators included clear explanations and feedback during examinations [18,48]. Barriers included insufficient guidance and lack of SDM training for HCPs [30,55]. “Terminology used by clinicians” was a facilitator when HCPs used layman’s terms and adjusted explanations to the patient’s health literacy [18,33,48]. Barriers included use of advanced medical terms [18,48]. “Non-verbal communication” was reported as a barrier by HCPs, with reliance on non-verbal cues sometimes leading to misinterpretation of patient consent or discomfort [30]. “Language barriers” were both a barrier and facilitator. Patient barriers included lack of in-depth discussion in a non-native language and poor interpreter services [69]. HCPs also saw language barriers as restricting SDM use [23]. A facilitator was linguistic compatibility between provider and patient [69].

#### 3.6.4. Knowledge of the application and use of shared decision-making

This theme, primarily represented by HCPs, included four subthemes: “familiarity with SDM,” “assumed efficacy vs informed choices,” “timing of shared decision-making,” and “clinical situation.” The subtheme “familiarity with SDM” was related to barriers such as a lack of comprehensive SDM courses [23], and facilitators included pre-existing knowledge of SDM [23,24]. The subtheme “assumed efficacy vs informed choices” was only reported as a barrier, where HCPs made management decisions based on personal preferences rather than informed patient choices [30,42,55,78] and used a patient-centred approach to gain compliance with their expert recommendations [78]. The subtheme “timing of SDM” was both a facilitator and barrier. Facilitators included more complex treatment decisions, where highrisk treatments promoted SDM, while low-risk decisions favoured informed consent. Barriers included HCPs relying on patients to refuse treatment rather than seeking consent and using intuition to decide when to apply SDM [1,24,30]. The subtheme “clinical situation” was related to barriers such as decision aids being too difficult to use [42,78] and not suitable for some clinical situations [30,72]. However, decision aids were also considered a facilitator as they helped ensure all management options were considered [72].

#### 3.6.5. Skills

This theme, represented by both patients and HCPs, included two subthemes: “SDM training” and “therapeutic alliance” [7,8,11,21,33,52,71]. Formal SDM training was considered a facilitator by both HCPs [7,8,19] and patients [7,8,11]. However, a lack of formal SDM training was reported as a barrier by HCPs [11,23], and patients also identified it as a barrier when SDM training did not lead to improved treatment satisfaction or effective implementation [11,64]. The subtheme “therapeutic alliance” was primarily considered a facilitator by both patients [7,8,11,21,33,52] and HCPs [7,8,30,33,71]. Factors contributing to a good therapeutic alliance reported by patients included feeling understood, being involved in SDM, having a sustained relationship with the HCP, and the HCP’s competence. For HCPs, positive feelings towards patients, SDM training, and trust were key factors. Conversely, the lack of a therapeutic alliance was seen as a barrier by both patients [11,63] and HCPs [30]. HCPs noted barriers such as not providing enough information during consultations and feeling underconfident when addressing psychological aspects of pain.

### 3.7. Interactional dynamics

#### 3.7.1. Decision aid

This theme, represented by both patients and HCPs, included four subthemes: “facilitating preparedness to participate in SDM using decision aids,” “decision aids features and user experience,” “checklist diagnosing,” and “wording of decision aid” [39,41,44,47,72]. Decision aids were generally considered facilitators of SDM. HCP facilitators included a functional approach with treatment options and psychological support [72]. Barriers for HCPs were related to decision aids hindering personal contact and being time-consuming [48,68]. Patient facilitators included improved consultations, knowledge of treatment options, and better trust in HCPs, while older patients found decision aids both a barrier and facilitator [39,41,44,47,68]. “Facilitating preparedness to participate in SDM using decision aids” was a facilitator for patients, as decision aids reduced decisional conflict and enhanced self-efficacy [15,31]. HCPs also noted patients becoming more psychologically ready for SDM [68]. “Decision aid features and user experience” were facilitators when decision aids had user-friendly designs, tracking features, and strong communication tools like SMS and videos [44,47]. “Checklist diagnosing” was a facilitator for HCPs in confirming all options were considered and supporting diagnostic thinking [42,72]. Barriers included the decision aid being too generic to account for patient differences [42]. For patients, trusting the HCP and asking relevant questions were facilitators [72]. “Wording of decision aids” was a barrier for both patients and HCPs. Patient barriers included closed questions restricting open discussion [72], while HCPs reported awkward phrasing or misaligned questions causing misunderstandings [72].

#### 3.7.2. Decision dynamics: Navigating patient-HCP roles, values, and power

This theme, represented by both HCPs and patients, included five subthemes: “decision characteristics,” “sharing responsibility,” “patient wants paternalistic decision-making,” “asking patients about their preferred role in decision-making,” and “asking patients about values/clarifying values” [11,18,69,72]. The subtheme “decision characteristics” was only represented by patients and was both a barrier and a facilitator. Barriers included high-stakes decisions where patients sought advice from relatives, though this was also a facilitator to SDM [69]. The subtheme “sharing responsibility” was represented by both HCPs and patients. Patient facilitators included involvement in decision-making, leading to increased motivation, understanding, and trust [24,33]. Barriers for patients included feeling disrespected or disbelieved by HCPs [21]. HCP facilitators were related to patient empowerment, and decision aids were found to encourage patient responsibility in decision-making [42,78]. The subtheme “patients want paternalistic decision-making” was considered a barrier by both patients and HCPs. HCP barriers included patients actively refusing to participate in SDM due to unfamiliarity with their rights or a preference for a provider-driven approach [8,23,52,68]. Patient barriers included believing the HCP should make decisions due to their expertise [11,18,69,72]. The subtheme “asking patients about their preferred role in decisionmaking” was reported by both patients and HCPs. Patient facilitators included increased motivation and trust in the HCP [24,68]. Barriers for patients included not being asked about their preferred role [63], while HCPs saw it as a barrier because it could imply a lack of knowledge or experience [68]. The subtheme “asking patients about values/clarifying values” was reported by both HCPs and patients. Facilitators included revisiting the patient’s beliefs during treatment, which engaged and motivated patients [63,78]. Barriers included not asking patients about their values and offering default treatment options instead [78].

### 3.8. System-level factors

#### 3.8.1. Cost-beneficial

The cost-beneficial aspect of SDM was a barrier reported by both HCPs [23,42] and patients [11,42,64]. HCP perceived barriers included lack of effectiveness and added value, risk of losing patients wanting passive care, and increased costs and time. Patient-related barriers were increased time and/or costs. However, HCPs also reported decision aids as a facilitating SDM intervention due to their reduction of workload and time [42]. Both patients and HCPs reported that decision aids were believed to reduce costs by improving decision accuracy in the first consultation and decreasing incorrect decisions, thereby reducing costs [68].

#### 3.8.2. Healthcare system organization

This theme, represented by both patients and HCPs, included six subthemes: “characteristics of healthcare setting,” “healthcare process expectancy,” “lack of resources,” “lack of access,” “organizational constraints,” and “multidisciplinary care.” The subtheme “characteristics of healthcare setting” was both a barrier and facilitator. Facilitators included thorough assessments and a comfortable environment [18,24,72]. Barriers included referral negotiations, high costs, and a rushed environment [11,18,21]. HCPs considered clinic characteristics both a facilitator and barrier, with economic benefits sometimes discouraging SDM [23,68]. “Healthcare process expectancy” was a barrier, with both patients and HCPs believing SDM wouldn’t improve healthcare. HCP barriers included concerns about confusion, patient loss, and limited specialized services [68,72,78]. “Lack of resources,” “lack of access,” and “organizational constraints” were barriers for both HCPs and patients. HCPs reported financial constraints, lack of specialists, and heavy workloads [23], while both groups noted limited access to services and SDM training [11,23,72]. “Organizational constraints” were both a barrier and facilitator, depending on the attitudes of supervisors [23,24]. “Multidisciplinary care,” reported only by patients, was both a barrier and facilitator. Facilitators included multidisciplinary care with expert input [11,33], while a barrier was difficulty in forming partnerships within the team [33].

### 3.9. Theoretical domains framework analysis

The outcome of the inductive analysis was mapped into the TDF utilising the predetermined criteria, as illustrated in **Table 2**. Across all contextual levels, key barriers and facilitators included knowledge, skills, and beliefs about consequences, and environmental context and resources. For patients, the most dominant facilitator besides the abovementioned was emotions. For HCPs, the most dominant facilitators besides the abovementioned were social/professional roles. Observers reported facilitators in the domains of knowledge, skills, and memory, attention, and decision processes. The most dominant domains identified as barriers for patients did not differ from the abovementioned. For HCPs, the most dominant barriers besides the abovementioned were social/professional roles and beliefs about capabilities. Observers reported barriers in the domains of knowledge, skills, and beliefs about consequences. See **Figure 3** for the number of studies identifying implementation barriers and facilitators to SDM related to the TDF domains for patients, healthcare professionals (HCPs), both groups, and observers.

**FIGURE 3.**
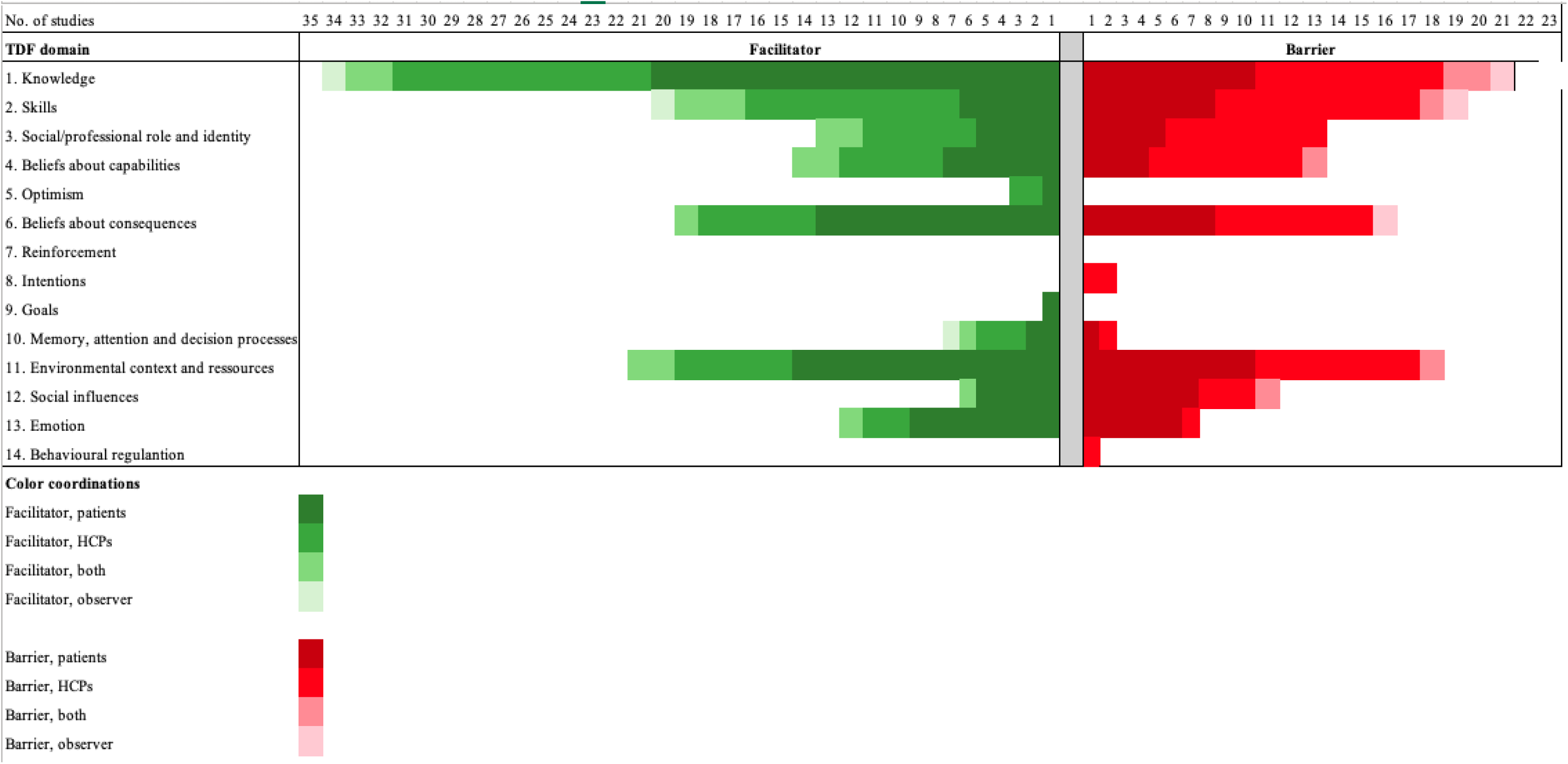
NUMBER OF STUDIES IDENTIFYING TDF BARRIERS AND FACILIATORS IN RELATION TO SDM.

**TABLE 1.** IN– AND EXCLUSION CRITERIA.

|  | Inclusion | Exclusion |
| --- | --- | --- |
| **Participants** | Healthcare providers<br>Relatives (proxy decision makers, caregivers, support persons?)<br>Adults aged 18 or above diagnosed with chronic musculoskeletal pain. | All studies not concerning individuals with a diagnosis of chronic musculoskeletal pain |
| **Interventions** | All SDM-related interventions across all clinical settings | All non-SDM-related interventions in any setting, including hypothetical SDM and non-clinical settings. |
| **Comparisons** | All comparisons | N/A |
| **Outcomes** | Barriers and/or facilitators of SDM within the healthcare system in terms of the management of CMP | N/A |
| **Studies** | All English-language study types with original data published in a peer-reviewed journal | All study types without original data and unpublished results. |

**TABLE 2.** MAPPING OF TDF DOMAINS.

| Shared Decision-Making and Chronic Musculoskeletal Pain |  |  |  |  |  |
| --- | --- | --- | --- | --- | --- |
| Stakeholders | HCP | Patient | Institutional | Organisational | System-level |
| **1. Knowledge** |  |  |  |  |  |
| Barriers | - Does not have enough knowledge on SDM (7)<br>- Aware of SDM but does not apply it (17), (14)<br>- Lack of SDM courses (7), (58)<br>- Assuming information giving is not necessary or will confuse patients (8)<br>- Under confidence in knowledge on SDM to perform it (9), (7)<br>- Insecure if explanations were sufficient (17)<br>- Feeling pressured to provide a diagnosis from patients (9)<br>- Differing explanations of pain in between HCPs (15)<br>- Diverging belief of pain origin (9)<br>- Managing unrealistic expectations of care from patients (9)<br>- Does not understand and believe DAs improves patient care (9), (58), (13)<br>- Inappropriate wording of DAs (14)<br>- Lack of evaluation of SDM interventions (58)<br>- Influence of other (18)<br>- HCP does not believe in SDM (17), (8)<br>- Incorrect use of SDM (8)<br>- Wants paternalistic care/does not want to engage in SDM (2), (58), (7) | - HCPs needs training to improve knowledge and facilitate SDM (4)<br>- HCP seems under confident in presenting phycological aspects of pain (9)<br>- Belief of pain origin (9)<br>- Non-credible treatment delivered by a wrong practitioner non-congruent with pain belief (12)<br>- HCP using medical terms to explain findings (19), (10)<br>- Being too knowledgeable to receive new treatment options (2)<br>- Insufficient explanation of pain, e.g. inconsistent explanations (10), (19), (14)<br>- No new advice given (11)<br>- Fear of pain stigma (9), (4), (12)<br>- Wants paternalistic care/does not want to engage in SDM (10), (14), (20), (4)<br>- Receiving a diagnosis (12)<br>- Lack of individualised care (10), (19), (11) | OBSERVER<br>- Not meeting patients expectations and preferences (19) |  |  |
| Facilitators | - SDM training for HCPs improved | - SDM training for HCPs | OBSERVER |  |  |
| Stakeholders | HCP | Patient | Institutional | Organisational | System-level |
|  | knowledge on SDM (2), (6), (18)<br>- Understanding what SDM entails (8), (17), (58)<br>- Has enough knowledge on SDM to perform it (7)<br>- Patient education (16), (58)<br>- Understands and believes DAs improves patients care (13), (58)<br>- Observing patients benefit from SDM (7)<br>- Influence of others (18)<br>- Using SDM in high-risk treatments (8)<br>- Using DAs as a checklist for thorough assessments (14), (13)<br>- Agreeing on belief of pain origin (9) | improved knowledge on SDM (2), (4),<br>- Support from HCPs (12), (11), (58)<br>- Patient education (3), (17), (18), (10), (12), (4), (16), (23)<br>- In-depth explanation of treatment choices (20), (19), (10)<br>- HCP communicating findings in layman's terms (19), (10), (11)<br>- Belief of pain origin (9)<br>- Credible treatment delivered by the right practitioner congruent with pain belief (12)<br>- Individualised care (10), (11), (4), (12), (14)<br>- Explanations from a HCP with clinical expertise within their field (11)<br>- Thorough assessment fosters better foundation for SDM (10)<br>- Being involved in the decision making (58)<br>- Receiving a diagnosis (12)<br>- DA's makes consultations more efficient, thorough and provides patient education (14), (22), (23), (24), (25), (58), (26), (27)<br>- Features of DAs enhancing SDM (16), (23)<br>- HCP aware of cultural differences and cultural competent (20), (24)<br>%21 | - HCP delivering clear information and ruling out serious disease (19) |  |  |
| **2. Skills** |  |  |  |  |  |
| Barriers | - Lack of SDM after formal SDM | - Lack of formal SDM training for | OBSERVER |  |  |
| Stakeholders | HCP | Patient | Institutional | Organisational | System-level |
|  | training (3), (5)<br>- Lack of formal SDM training resulting in lack of SDM skills (7), (58), (4)<br>- Lack of confidence in own skills (17), (9)<br>- Not having a therapeutic alliance (8)<br>- Lack of therapeutic alliance due to under confidence in skills and anxiety performing SDM on patients with a psychological overlay (9), (7)<br>- Managing unrealistic expectations from patients (9)<br>- SDM modulated to own personal preferences (17), (8)<br>- Threat to autonomy (13)<br>- Lack of communication skills (7), (19)<br>- Not applicable given the clinical situation (13), (8) | HCPs (4)<br>- Lack of therapeutic alliance (4)<br>- Multidisciplinary care: not being able to develop a proper HCP-patient relationship (11)<br>- Lack of HCP communication skills (10), (14)<br>- Language barriers restricting use of SDM (20)<br>- Group rehabilitation not taking individualized care into account (10), (11), (19)<br>- Younger age ease the use of DAs (22) | - Communication skills, e.g. being patient-centered (19) |  |  |
| Facilitators | - Formal SDM training for HCPs (1), (2), (3), (18), (7)<br>- Building a strong therapeutic alliance (1), (6), (8)<br>- Using DAs to guide SDM (13), (58)<br>- Adjusting SDM to the clinical situation (17)<br>- Revisiting pain beliefs and applying SDM throughout course of treatment (9)<br>- Having good communication skills (19)<br>- Expressing interest in the patients (19) | - Formal SDM training for HCPs (1), (2), (4)<br>- Building a strong therapeutic alliance (2), (11), (4)<br>- Competence of the doctor (4)<br>- Multidisciplinary care and care provided from HCPs with expertise in different fields (11)<br>- Receiving a thorough examination (10)<br>- Revisiting pain beliefs and re applying SDM throughout course of treatment (9)<br>- Good communication skills of HCP (10), (11)<br>- Linguistic compatibility when | OBSERVER<br>- Communication skills, e.g. effective reassurance (19) |  |  |
| Stakeholders | HCP | Patient | Institutional | Organisational | System-level |
|  |  | language barriers are present (20)<br>- HCP characteristics and skills (12), (11)<br>- Older age challenges the use of Das (22) |  |  |  |
| **3. Social/professional role and identity** |  |  |  |  |  |
| Barriers | - Dissonance in in beliefs of pain origin (9)<br>- Power imbalance in partnership (17)<br>- Threat to autonomy (13)<br>- Believing patients are not capable of making a choice (17)<br>- Influence of others restricting use of SDM (18)<br>- Choosing own preferred treatment rather than SDM (17), (8), (14)<br>- Using SDM can be misunderstood as lack of knowledge (58)<br>- Characteristics of patients, e.g. fear avoidance, lack of motivation, lack of adherence (7)<br>- Patient wants paternalistic care (20) | - Power imbalance in the partnership (4)<br>- Pain stigma (4)<br>- Different HCPs at every encounter (11)<br>- Patient wants paternalistic care (14) |  |  |  |
| Facilitators | - SDM facilitated having less negative feelings towards patients (2)<br>- Building rapport enhanced partnership and SDM (8)<br>- A good partnership enhances mangitude of interaction and adoption of SDM (11) | - A good partnership enhances magnitude of interaction and adoption of SDM (11), (18)<br>- HCP's supporting, advocating for patients and negotiating the system (12), (11)<br>- Competence of HCP (4)<br>- Characteristics of HCP, e.g. |  |  |  |
| Stakeholders | HCP | Patient | Institutional | Organisational | System-level |
|  | - Observing patients benefit from SDM (7)<br>- Influence of others facilitating use of SDM (18)<br>- Enhanced focus on SDM when using risky treatments (8)<br>- Clear policy information (13)<br>- Trust (8), (19) | - empathetic interactions and emotional support (20), (10), (12), (19), (11)<br>- Having a sustained relationship with a HCP (11)<br>- Trust (20), (10), (4) |  |  |  |
| **4. Beliefs about capabilities** |  |  |  |  |  |
| Barriers | - Lack of confidence in applying SDM on pt with psychological overlay (9), (7)<br>- Feeling pressured to provide a diagnosis (9)<br>- Wanting to give the patient a diagnosis despite not being sure (17)<br>- Believing SDM is not useful and the intention to use it (13)<br>- Not feeling competent performing SDM (9)<br>- Fear of DA's does not work correctly (13) or the wording is inappropriate (14)<br>- Patients does not want to participate in SDM despite being offered (2), (14)<br>- Clinician beliefs patients are not capable to perform SDM, e.g. low level of education (58) | - Believing SDM does not make the patient feel good (18)<br>- Being offered similar treatment without showing effectiveness (11)<br>- Lack of health literacy to direct treatment pathway (4) |  |  |  |
| Facilitators | - Improved quality of SDM after training (1)<br>- Being able to realistically estimate patients' satisfaction (2)<br>- Believing SDM is useful and the intention to use it (13) | - Improved quality of SDM after training of HCPs (1)<br>- Systematic active approach to monitoring and evaluation of symptoms (12)<br>- Believing SDM makes the |  |  |  |
| Stakeholders | HCP | Patient | Institutional | Organisational | System-level |
|  | - Believing DA's are useful as a supplementary checklist for a thorough exam (14) and to raise psychological issues (14)<br>- Believing patients autonomy and valuing their rights should be taken into consideration (58)<br>- Features of the DA facilitates SDM (16) | patient feel good (18)<br>- Wanting to be included in clinical decision-making (18), (11)<br>- DAs facilitates the partnership (25)<br>- Features of the DA facilitates SDM (16) |  |  |  |
| **. Optimism** |  |  |  |  |  |
| Barriers |  |  |  |  |  |
| Facilitators | - Feeling optimistic using SDM (7)<br>- Belief that patients benefits from using SDM (13) | - Feeling listened to, understood and contained by HCPs (2) |  |  |  |
| **6. Beliefs about consequences** |  |  |  |  |  |
| Barriers | - Different beliefs of pain origin (9)<br>- Discussing psychosocial aspects of pain (9)<br>- Managing patients' unrealistic expectations of care (9)<br>- Belief that SDM does not improve patient care (13)<br>- Believing patients are not capable of making er choice (17)<br>- Choosing own preferred treatment rather than SDM (17), (8), (14)<br>- Patients wants paternalistic care (9), (2), (10), (20), (4)<br>- Patient characteristics (7), (2)<br>- Not taking enough time to explain course of treatment (8)<br>- DAs hampers personal contact to perform SDM (13) | - No new advice during consultations (19)<br>- Perception of own health at a certain age (9)<br>- Pain stigma and disbelief from HCPs (4), (19)<br>- SDM exacerbates pain (18)<br>- Receiving multidisciplinary care makes it difficult to develop a proper partnership (11)<br>- Not being offered involvement in decision making (10)<br>- Not providing individualised care taking daily life situations into account (19)<br>- Discussing psychosocial aspects of pain (4), (19)<br>- Cultural differences in the HCP-patient relationship (21) | OBSERVER<br>- Mismatch in HCP and patient agenda (19) |  |  |
| Facilitators | - Perceived usefulness, benefit | - Systematic active approach to |  |  |  |
| Stakeholders | HCP | Patient | Institutional | Organisational | System-level |
|  | and intention to use SDM (13), (15)<br>- Using SDM in treatments considered risky (8)<br>- Patients taking responsibility for their own health (9)<br>- Continuously revisiting beliefs of pain performing SDM (9)<br>- Using DA to raise psychological issues (14)<br>- Using DAs improves assessment of patients (13) | monitoring and evaluation of symptoms of ongoing therapeutic relationship (12)<br>- New advice during consultations (19)<br>- Perception of own health at a certain age (9)<br>- SDM makes the patient feel better about themselves (18)<br>- Receiving multidisciplinary care facilitates SDM (11)<br>- Taking responsibility for own care and contributing to shared responsibility (11)<br>- Continuously revisiting beliefs of pain performing SDM (9)<br>- Well explained treatment options facilitated SDM (10)<br>- Individualised care (10), (11), (4), (12), (4)<br>- Being open and honest throughout the consultation fosters a better relationship (4)<br>- Using DAs leads to more thorough consultation and better information on treatment options (14), (22), (24), (25)<br>- HCPs addressing psychological concerns (14)<br>- Turning to friends/acquainted with medical knowledge for advice (20)<br>- Cultural difference: having cultural competent provider (20), (24)<br>- Cultural differences in the HCP-patient relationship (21) |  |  |  |
| **7. Reinforcement** |  |  |  |  |  |
| Stakeholders | HCP | Patient | Institutional | Organisational | System-level |
| Barriers |  |  |  |  |  |
| Facilitators |  |  |  |  |  |
| **8. Intentions** |  |  |  |  |  |
| Barriers | - Not offering the patient the option of choosing between treatment options (17), (7)<br>- Choosing treatment despite asking about preferences (17) |  |  |  |  |
| Facilitators |  |  |  |  |  |
| **9. Goals** |  |  |  |  |  |
| Barriers |  |  |  |  |  |
| Facilitators |  | - Identifying and setting individual goals with HCP (11) |  |  |  |
| **10. Memory, attention, and decision processes** |  |  |  |  |  |
| Barriers | - Fear of DA not working correctly (13) | - Being offered same treatment options that previously has shown no effect (11) |  |  |  |
| Facilitators | - Building rapport to choose treatment direction (8)<br>- Using DA as a checklist to provide all suitable options and discuss various aspects of pain (14)<br>- Revisiting patients' beliefs regularly during treatment (9) | - Individualised care (17), (10), (11) | OBSERVER<br>- Communicating clear results for the patient to draw conclusions on their own (19) |  |  |
| **11. Environmental context and resources** |  |  |  |  |  |
| Barriers | - Lack of training offers to improve knowledge and skills (7), (4) | - Negative HCP-patient partnership (4) |  |  |  |
| Stakeholders | HCP | Patient | Institutional | Organisational | System-level |
|  | - Lack of time (11), (Rashida), (4), (7)<br>- Constrains to perform SDM in existing services (11)<br>- Not cost-beneficial (7), (13), (58)<br>- Breaking trust in authority and appearing insecure (8)<br>- Having economic hidden agendas (58)<br>- Characteristics of clinics, e.g. lack of space, equipment, staff shortage (7).<br>- Loss of potential income and increased time restraining use of SDM (7), (13)<br>- Lack of access to refer to specialised services due to geographic location or long waiting lists (14)<br>- Lack of specialists per capita (58)<br>- Clinic policy/behaviour prioritising some treatments over others not using SDM (7)<br>- Influence of colleagues not wanting to engage in SDM (18)<br>- Applying SDM by own preferences (7) and clinical situation (13)<br>- Length of pain of the patient and length of patient-HCP partnership (9)<br>- DA hampering personal contact (13), (14) and increases time (58)<br>Characteristics of patients (7) | - Not cost-beneficial (4), (5)<br>- Healthcare system characteristics, e.g. negotiation of tests, referrals and treatments, costs of care, rescheduling appointments (12), (4), (10), (11)<br>- Lack of time (4), (11), (18), (11) and HCPs being rushed (10), (11).<br>- Long waiting times (10)<br>- Rushed environment at GP with limited time (11)<br>- Lack of access to specialised services due to geographic location or long waiting lists (14)<br>- Influence of colleagues wanting to engage in SDM (18)<br>- Lack of high quality interpreter services when language barriers are present (20)<br>- Length of pain of the patient and length of patient-HCP partnership (9)<br>- Older age eased instructions to SDM/DA's (22)<br>- DA restricting personal contact (14) |  |  |  |
| Facilitators | - Cost-beneficial (58), (13)<br>- Observing patients benefit from SDM (7).<br>- Characteristics of the clinic, e.g. | - Training of HCP skills (4)<br>- Multidisciplinary care (4)<br>- Advocacy of family doctors to negotiate the system (12) |  |  |  |
| Stakeholders | HCP | Patient | Institutional | Organisational | System-level |
|  | equipment, space, collaborative colleagues (7)<br>- Length of pain of the patient and length of patient-HCP partnership (9) | - Positive HCP-patient relationship (4)<br>- Time (3)<br>- Using DA to be more time-effective and thorough (14), (25), (58)<br>- Cost-beneficial (58)<br>- Healthcare system characteristics, e.g. thorough assessment, homely environment, structural conditions (10), (18), (11)<br>- Pain clinics offers more time and a more holistic approach (11)<br>- Cultural factors facilitating trust in ones doctor (21)<br>- Characteristics of HCP enhances SDM (12)<br>- Length of pain of the patient and length of patient-HCP partnership (9)<br>- Younger age eased instructions to SDM/DA's (22)<br>- Features of DA (16), (23) |  |  |  |
| **12. Social influences** |  |  |  |  |  |
| Barriers | - Non-acceptance of psychological aspects of pain (9)<br>- Preferring passive care (7)<br>- Cultural differences (7)<br>- Length of partnership (9)<br>- Patient characteristic (58), (7), (4) | - Misalignment in belief of pain with HCP (9)<br>- Power imbalance between patient and HCP (4), (9), (12), (14), (19),<br>- Perception of own age-related health (9)<br>- Length of partnership (9)<br>- Turning to relatives for decision making (20)<br>- Turning to others at high-stake |  |  |  |
| Stakeholders | HCP | Patient | Institutional | Organisational | System-level |
|  |  | decisions (20) |  |  |  |
| Facilitators | - Clinicians wants clinical judgment trusted without (9)<br>- Culture (21)<br>- Length of partnership (9) | - Aligning belief of pain with HCP (9)<br>- Perception of own age related health (9)<br>- Length of partnership (9)<br>- Cultural competent HCPs (24), (20)<br>- Turning to relatives for decision making (20)<br>- Turning to others at high-stake decisions (20) |  |  |  |
| **13. Emotion** |  |  |  |  |  |
| Barriers | - Dissonance in belief of pain origin reducing legitimization of pain and trust in the relationship (9)<br>- Abruptness and not being empathetic (10) | - Having a sustained relationship with a HCP (9)<br>- Not receiving emotional support from the HCP (9)<br>- Strong beliefs of pain origin (9)<br>- Fear of being seen as a hypochondria (4), (12)<br>- SDM exacerbated pain and made pt feel anxious (18)<br>- Not considering how MSK pain affects ones life (19)<br>- Receiving a diagnosis is considered labelling (12)<br>- Being put off with psychological explanations (9), (19) |  |  |  |
| Facilitators | - Having fewer negative feelings toward the patients (2) | - Emotional support and pain legitimization from HCPs (2), (9), (20), (10), (4), (19)<br>- Having a sustained relationship with a HCP (9)<br>- Strong beliefs of pain origin (9)<br>- SDM making the patients feel better (18)<br>- Receiving a diagnosis was considered as a relief ruling out |  |  |  |
| Stakeholders | HCP | Patient | Institutional | Organisational | System-level |
|  |  | serious disease (12)<br>- Exploring psychological aspects of pain (14), (9) |  |  |  |
| **14. Behavioural regulation** |  |  |  |  |  |
| Barriers | - Making decision based on personal preferences rather than mutual agreements (17) |  |  |  |  |
| Facilitators |  |  |  |  |  |

## Discussion

This scoping review identified facilitators and barriers to the implementation and use of SDM in the management of chronic musculoskeletal pain across different contextual levels. Our findings highlight the complexity of utilising SDM in clinical practice, highlighting the need for coordinated implementation efforts at system, clinician, and patient levels. For patients, key facilitators included HCP training in SDM, therapeutic alliance, trust, empathy, and knowledge of condition and treatment options. Individualized care and decision aids also supported patient participation. Patient barriers included poor were primarily related to communication, lack of individualized care, and insufficient explanation of the condition and treatment options. Additionally, time constraints and healthcare system challenges hindered SDM. For HCPs, facilitators included formal SDM training, communication skills, and a positive attitude towards SDM. Barriers reported by HCPs included concerns about autonomy, lack of familiarity and training with SDM, time constraints, and challenges in applying SDM in certain clinical situations. This review is the first to comprehensively map barriers and facilitators specific to musculoskeletal pain across all sectors and countries. Our inductive analysis revealed previously underexplored themes and offered deeper insight into how context shapes SDM implementation in practice.

In recent years, extensive research has been conducted to investigate barriers and facilitators to both implementation and use of SDM across different healthcare systems targeting different health conditions [10,20,29,32,36,43,45,50,62,65,67,70,73,81,84]. Our review builds on this literature but addresses the underexplored context of chronic musculoskeletal pain, which poses distinct challenges due to high rates of diagnostic uncertainty, overlapping psychosocial factors, long-term fluctuating symptoms, and fragmented care pathways. These complexities make it more difficult to meaningfully individualize care, sustain therapeutic relationships, and integrate SDM into routine decision-making, despite its potential benefits. This echoes previous findings of an “intention–action gap,” where clinicians support SDM conceptually but fail to implement it consistently due to time and resource constraints [45]. Similarly, our study emphasizes the recurring challenge of time constraints as a barrier to SDM in chronic musculoskeletal care, a finding also supported by multiple reviews across different healthcare settings [10,45,50,65,84]. Previous research indicates that while SDM is often perceived to be more time-consuming, there is little evidence that it significantly increases consultation length [40,51,75,82]. This finding also emerged from our study, challenging the clinical assumption that SDM inherently requires more time compared to standard consultations. Instead, access to resources and training appears more critical for effective implementation than consultation length. These barriers must be addressed to make SDM sustainable in musculoskeletal care.

For SDM to be integrated effectively within health systems, a systematic implementation plan that involves addressing these misconceptions and collaborating closely with internal stakeholders is crucial [14,40]. Gravel et al. 2006 and Légaré et al. 2008 noted that a lack of training, communication skills, and decision support tools frequently limit clinicians’ ability to operationalize SDM in everyday practice [36,50]. To combat this, SDM training has gained interest and has now been introduced in undergraduate medical programs [22,85]. Despite promising results, research has suggested a shift from passive and theory-based learning to active and practice-based learning to further facilitate the implementation and appropriate use of SDM [16,61]. Future work should explore how SDM training can be tailored to individual musculoskeletal conditions, healthcare settings, and provider needs.

Another important factor impacting the use of SDM was the lack of patients’ knowledge and information about their musculoskeletal condition. Across the included studies, patients identified educational materials and adequate plain language explanations of their conditions as significant facilitators for increasing their knowledge. The development of decision aids has helped meet the need for comprehensive information, enabling informed patient choices and facilitating SDM [76], which were supported by our review. We found that effective SDM in chronic musculoskeletal care hinges not only on technical skills or decision aids, but on the ability to build trust and navigate uncertainty. Patients emphasized the need for accessible explanations of their condition and treatment options, ideally grounded in their personal context and delivered with empathy. Decision aids, while helpful, were only effective when aligned with the patient’s understanding of their pain and used as part of a relational, not transactional, conversation. Scholl et al. 2018 highlighted organizational factors such as leadership, workflow integration, and policies as key influencers of SDM uptake [73], and we observed similar barriers in the context of musculoskeletal care, but found they were further complicated by condition-specific issues like diagnostic uncertainty and incoherent care pathways.

Légaré et al. 2018 emphasized the need for broader implementation strategies, including audit, feedback, and institutional support [49]. Ubbink et al. 2024 conducted a review of barriers and facilitators to SDM for emergency-departments and found that implementation tools such as checklists and decision aids were more effective when accompanied by local champions and integrated into electronic health records [81]. Similarly, Waddell et al. 2021 showed that barriers extend across all domains of the TDF including barriers and facilitators across individual, organizational, and system levels [84]. The study found that the SDM process often was reduced to a communication technique rather than a truly shared deliberative process [84]. Our study supports these conclusions but contributes musculoskeletal-specific insights. For example, we found that passive dissemination of decision aids was rarely sufficient; effective implementation required adaptations to existing care models. Francis et al. 2024 found that supported decision-making requires not only informational support but also relational scaffolding, e.g., trust-building, legitimization of patient narratives, and attention to power asymmetries [32]. Similarly, Pel-Littel et al. 2021 highlighted the need for personalization, particularly for older adults managing multiple conditions, where SDM must be sensitive to fluctuating goals and capacities [65]. These insights translate directly to musculoskeletal care, where patients often cycle through phases of flare-up and remission and shifting priorities over time. This underscores the need for SDM approaches in musculoskeletal care to support ongoing, co-produced care planning that can adapt to evolving patient needs. In line with this, the therapeutic alliance between patients and HCPs is a critical factor for the use and implementation of SDM [86], also support by our review. Our findings show that patient and HCP characteristics, attitudes, perspectives towards SDM, trust, and communication, are interrelated and influence the therapeutic alliance. Trust is a key facilitator of SDM, shaped by both parties’ characteristics and communication skills, where poor communication can breed uncertainty and distrust. Decision dynamics, such as patient preference for paternalistic care, and HCP attitudes towards SDM varying by decision risk, also influence the therapeutic alliance. Importantly, our review reveals that these dynamics are particularly complex in musculoskeletal care, where the perceived legitimacy of pain, uncertainty around causation, and long-term provider relationships can either strengthen or undermine the foundation needed for SDM. These interconnected dynamics underscore the need for future SDM research in musculoskeletal care to focus on how therapeutic alliances can be fostered and sustained amid diagnostic uncertainty, fluctuating symptoms, and diverse patient and HCP preferences.

A key strength of this scoping review is its comprehensive synthesis of both conceptual and empirical literature on SDM implementation in chronic musculoskeletal pain. By combining previously established taxonomies and an inductive thematic analysis with the TDF, we offer a nuanced understanding of multilevel barriers and facilitators, incorporating diverse perspectives from patients, healthcare professionals, and system-level factors across various study designs and contexts. However, limitations include the exclusion of non-English studies, a focus on breadth rather than depth, reliance on self-reported data, and the inclusion of studies on SDM in broader contexts beyond chronic musculoskeletal pain. Despite these, this review provides a valuable foundation for future SDM research and practice in chronic pain care.

## Conclusion

This study identified key facilitators and barriers to the implementation of SDM in managing chronic musculoskeletal pain. Facilitators at the patient level included HCPs trained in SDM, strong patient-HCP relationships, empathetic care, education on the condition and treatment options, and decision aids. For HCPs, facilitators included formal SDM training, effective communication, positive attitudes, and the use of decision aids. Barriers for patients involved communication issues, lack of individualized care, insufficient knowledge, preferences for paternalistic decision-making, and time-related challenges. For HCPs, barriers included attitudes towards SDM, perceived threats to autonomy, lack of confidence, limited knowledge, time constraints, and difficulties applying SDM in clinical practice. Our TDF analysis highlighted that knowledge, skills, beliefs, and environmental context were common barriers and facilitators. This study underscores existing knowledge gaps, offering valuable insights for future research to optimize SDM implementation in chronic musculoskeletal pain management.

## Declarations

### Ethics approval and consent to participate

Not applicable.

## Consent for publication

Not applicable.

## Availability of data and materials

The datasets used and/or analysed during the current study are available from the corresponding author on reasonable request.

## Authors’ contributions

**Conceptualization:** All authors. **Methodology:** All authors. **Formal analysis:** LNA, KDL

**Investigation:** LNA, KDL. **Resources:** JFJ, MSR, and JLT. **Writing – Original Draft:** KDL, LNA. **Writing – Review and Editing:** All authors. **Visualization:** LNA and KDL.

