## Appendix 1 for "Implementation of Shared Decision-Making in the Management of Chronic Musculoskeletal Pain: *a scoping review*"

Search Strategy

Chronic Musculoskeletal Pain

### Databases and results

| Database | Interface | Result | Date | Result 1/12/24 |
| --- | --- | --- | --- | --- |
| PubMed | PubMed.gov | 1542 | 18.01.2023 | 170 |
| Embase | Embase.com | 1499 | 19.01.2023 |  |
| Cochrane Library | Wiley | 231 | 19.01.2023 |  |
| Scopus | Scopus.com | 972 | 20.01.2023 |  |
| PsycINFO | APA | 23 | 19.01.2023 |  |
| CINAHL with Fulltext | EBSCO | 654 | 19.01.2023 |  |
| PEDro | pedro.org.au | 109 | 19.01.2023 |  |
| All  After removal of duplicates with EndNote  After import to Covidence |  | 5030  3146 |  |  |

| Database | Interface | Result | Date | Result |
| --- | --- | --- | --- | --- |
| PubMed | PubMed.gov | 1542 | 09.05.2025 | 376 |
| Embase | Embase.com | 1499 | 09.05.2025 | 461 |
| Cochrane Library | Wiley | 231 | 09.05.2025 | 25 |
| Scopus | Scopus.com | 972 | 09.05.2025 | 165 |
| PsycINFO | APA | 23 | 09.05.2025 | 0 |
| CINAHL with Fulltext | EBSCO | 654 | 09.05.2025 | 372 |
| PEDro | pedro.org.au | 109 | 09.05.2025 | 10 |
| All  After removal of duplicates with EndNote  After import to Covidence |  | 5030  3146 |  | 1409 488 |

### PubMed

| Search | Query | Results |
| --- | --- | --- |
| #32 | Search: **((((((((((((("Musculoskeletal Pain"[Mesh]) OR ("Musculoskeletal Diseases"[Mesh])) OR ("Arthralgia"[Mesh])) OR ("Back Pain"[Mesh])) OR ("Facial Pain"[Mesh])) OR ("Headache"[Mesh])) OR ("Metatarsalgia"[Mesh])) OR ("Neck Pain"[Mesh])) OR ("Nociceptive Pain"[Mesh])) OR ("Pain, Intractable"[Mesh])) OR ("Pelvic Pain"[Mesh])) OR (musculoskeletal[Text Word] OR hip pain*[Text Word] OR knee pain*[Text Word] OR shoulder pain*[Text Word] OR neck pain*[Text Word] OR back pain*[Text Word] OR elbow pain*[Text Word] OR hand pain*[Text Word] OR headach*[Text Word] OR pelvic pain*[Text Word] OR facial pain*[Text Word] OR leg pain*[Text Word] OR bursitis[Text Word] OR tendin*[Text Word] OR tendon*[Text Word] OR myalgia[Text Word] OR arthralgia*[Text Word] OR joint pain*[Text Word] OR bone pain*[Text Word] OR ostalgia[Text Word] OR "Fractures, Bone"[Mesh] OR fracture*[Text Word] OR arthrit*[Text Word] OR rheumat*[Text Word] OR arthro*[Text Word] OR arm pain*[Text Word] OR muscle pain*[Text Word] OR "Soft Tissue Injuries"[Mesh] OR foot pain*[Text Word])) AND (("Chronic Pain"[Mesh]) OR ((chronic*[Text Word] OR recurr*[Text Word] OR continu*[Text Word] OR longstanding[Text Word] OR persist*[Text Word]) AND pain*[Text Word]))) AND ((((((((((((("Decision Making, Shared"[Mesh]) OR ("Decision Support Techniques"[Mesh:NoExp])) OR ("Decision Making"[Mesh:NoExp])) OR ("Decision Support Systems, Clinical"[Mesh])) OR ("Choice Behavior"[Mesh:NoExp])) OR (shared decision*[Text Word] OR sharing decision*[Text Word])) OR (informed decision*[Text Word] OR informed choice*[Text Word] OR decision aid*[Text Word])) OR ((share*[Text Word] OR sharing*[Text Word] OR informed*[Text Word]) AND (decision*[Text Word] OR deciding*[Text Word] OR choice*[Text Word]))) OR (decision making*[Text Word] OR decision support*[Text Word] OR choice behavio*[Text Word])) OR ((decision*[Title] OR choice*[Title]) AND (making*[Title] OR support*[Title] OR behavio*[Title]))) OR ("Patient Participation"[Mesh])) OR (patient participation*[Text Word] OR consumer participation*[Text Word] OR patient involvement*[Text Word] OR consumer involvement*[Text Word])) OR ((patient*[Title] OR consumer*[Title]) AND (involvement*[Title] OR involving*[Title] OR participation*[Title] OR participating*[Title] OR interact*[Title])))** Sort by: **Publication Date** | [1,542](https://pubmed.ncbi.nlm.nih.gov/?term=longquery105da8a3007dde12f9a2&sort=pubdate&size=20&ac=no) |
| #31 | Search: **(((((((((((("Decision Making, Shared"[Mesh]) OR ("Decision Support Techniques"[Mesh:NoExp])) OR ("Decision Making"[Mesh:NoExp])) OR ("Decision Support Systems, Clinical"[Mesh])) OR ("Choice Behavior"[Mesh:NoExp])) OR (shared decision*[Text Word] OR sharing decision*[Text Word])) OR (informed decision*[Text Word] OR informed choice*[Text Word] OR decision aid*[Text Word])) OR ((share*[Text Word] OR sharing*[Text Word] OR informed*[Text Word]) AND (decision*[Text Word] OR deciding*[Text Word] OR choice*[Text Word]))) OR (decision making*[Text Word] OR decision support*[Text Word] OR choice behavio*[Text Word])) OR ((decision*[Title] OR choice*[Title]) AND (making*[Title] OR support*[Title] OR behavio*[Title]))) OR ("Patient Participation"[Mesh])) OR (patient participation*[Text Word] OR consumer participation*[Text Word] OR patient involvement*[Text Word] OR consumer involvement*[Text Word])) OR ((patient*[Title] OR consumer*[Title]) AND (involvement*[Title] OR involving*[Title] OR participation*[Title] OR participating*[Title] OR interact*[Title]))** Sort by: **Publication Date** | [400,918](https://pubmed.ncbi.nlm.nih.gov/?term=%28%28%28%28%28%28%28%28%28%28%28%28%22Decision+Making%2C+Shared%22%5BMesh%5D%29+OR+%28%22Decision+Support+Techniques%22%5BMesh%3ANoExp%5D%29%29+OR+%28%22Decision+Making%22%5BMesh%3ANoExp%5D%29%29+OR+%28%22Decision+Support+Systems%2C+Clinical%22%5BMesh%5D%29%29+OR+%28%22Choice+Behavior%22%5BMesh%3ANoExp%5D%29%29+OR+%28shared+decision%2A%5BText+Word%5D+OR+sharing+decision%2A%5BText+Word%5D%29%29+OR+%28informed+decision%2A%5BText+Word%5D+OR+informed+choice%2A%5BText+Word%5D+OR+decision+aid%2A%5BText+Word%5D%29%29+OR+%28%28share%2A%5BText+Word%5D+OR+sharing%2A%5BText+Word%5D+OR+informed%2A%5BText+Word%5D%29+AND+%28decision%2A%5BText+Word%5D+OR+deciding%2A%5BText+Word%5D+OR+choice%2A%5BText+Word%5D%29%29%29+OR+%28decision+making%2A%5BText+Word%5D+OR+decision+support%2A%5BText+Word%5D+OR+choice+behavio%2A%5BText+Word%5D%29%29+OR+%28%28decision%2A%5BTitle%5D+OR+choice%2A%5BTitle%5D%29+AND+%28making%2A%5BTitle%5D+OR+support%2A%5BTitle%5D+OR+behavio%2A%5BTitle%5D%29%29%29+OR+%28%22Patient+Participation%22%5BMesh%5D%29%29+OR+%28patient+participation%2A%5BText+Word%5D+OR+consumer+participation%2A%5BText+Word%5D+OR+patient+involvement%2A%5BText+Word%5D+OR+consumer+involvement%2A%5BText+Word%5D%29%29+OR+%28%28patient%2A%5BTitle%5D+OR+consumer%2A%5BTitle%5D%29+AND+%28involvement%2A%5BTitle%5D+OR+involving%2A%5BTitle%5D+OR+participation%2A%5BTitle%5D+OR+participating%2A%5BTitle%5D+OR+interact%2A%5BTitle%5D%29%29&sort=pubdate&size=20&ac=no) |
| #30 | Search: **(patient*[Title] OR consumer*[Title]) AND (involvement*[Title] OR involving*[Title] OR participation*[Title] OR participating*[Title] OR interact*[Title])** Sort by: **Publication Date** | [18,529](https://pubmed.ncbi.nlm.nih.gov/?term=%28patient%2A%5BTitle%5D+OR+consumer%2A%5BTitle%5D%29+AND+%28involvement%2A%5BTitle%5D+OR+involving%2A%5BTitle%5D+OR+participation%2A%5BTitle%5D+OR+participating%2A%5BTitle%5D+OR+interact%2A%5BTitle%5D%29&sort=pubdate&size=20&ac=no) |
| #29 | Search: **patient participation*[Text Word] OR consumer participation*[Text Word] OR patient involvement*[Text Word] OR consumer involvement*[Text Word]** Sort by: **Publication Date** | [33,777](https://pubmed.ncbi.nlm.nih.gov/?term=patient+participation%2A%5BText+Word%5D+OR+consumer+participation%2A%5BText+Word%5D+OR+patient+involvement%2A%5BText+Word%5D+OR+consumer+involvement%2A%5BText+Word%5D&sort=pubdate&size=20&ac=no) |
| #28 | Search: **"Patient Participation"[Mesh]** Sort by: **Publication Date** | [29,096](https://pubmed.ncbi.nlm.nih.gov/?term=%22Patient+Participation%22%5BMesh%5D&sort=pubdate&size=20&ac=no) |
| #27 | Search: **(decision*[Title] OR choice*[Title]) AND (making*[Title] OR support*[Title] OR behavio*[Title])** Sort by: **Publication Date** | [43,910](https://pubmed.ncbi.nlm.nih.gov/?term=%28decision%2A%5BTitle%5D+OR+choice%2A%5BTitle%5D%29+AND+%28making%2A%5BTitle%5D+OR+support%2A%5BTitle%5D+OR+behavio%2A%5BTitle%5D%29&sort=pubdate&size=20&ac=no) |
| #26 | Search: **decision making*[Text Word] OR decision support*[Text Word] OR choice behavio*[Text Word]** Sort by: **Publication Date** | [328,959](https://pubmed.ncbi.nlm.nih.gov/?term=decision+making%2A%5BText+Word%5D+OR+decision+support%2A%5BText+Word%5D+OR+choice+behavio%2A%5BText+Word%5D&sort=pubdate&size=20&ac=no) |
| #25 | Search: **(share*[Text Word] OR sharing*[Text Word] OR informed*[Text Word]) AND (decision*[Text Word] OR deciding*[Text Word] OR choice*[Text Word])** Sort by: **Publication Date** | [72,648](https://pubmed.ncbi.nlm.nih.gov/?term=%28share%2A%5BText+Word%5D+OR+sharing%2A%5BText+Word%5D+OR+informed%2A%5BText+Word%5D%29+AND+%28decision%2A%5BText+Word%5D+OR+deciding%2A%5BText+Word%5D+OR+choice%2A%5BText+Word%5D%29&sort=pubdate&size=20&ac=no) |
| #24 | Search: **informed decision*[Text Word] OR informed choice*[Text Word] OR decision aid*[Text Word]** Sort by: **Publication Date** | [16,786](https://pubmed.ncbi.nlm.nih.gov/?term=informed+decision%2A%5BText+Word%5D+OR+informed+choice%2A%5BText+Word%5D+OR+decision+aid%2A%5BText+Word%5D&sort=pubdate&size=20&ac=no) |
| #23 | Search: **shared decision*[Text Word] OR sharing decision*[Text Word]** Sort by: **Publication Date** | [14,209](https://pubmed.ncbi.nlm.nih.gov/?term=shared+decision%2A%5BText+Word%5D+OR+sharing+decision%2A%5BText+Word%5D&sort=pubdate&size=20&ac=no) |
| #22 | Search: **"Choice Behavior"[Mesh:NoExp]** Sort by: **Publication Date** | [34,720](https://pubmed.ncbi.nlm.nih.gov/?term=%22Choice+Behavior%22%5BMesh%3ANoExp%5D&sort=pubdate&size=20&ac=no) |
| #21 | Search: **"Decision Support Systems, Clinical"[Mesh]** Sort by: **Publication Date** | [9,287](https://pubmed.ncbi.nlm.nih.gov/?term=%22Decision+Support+Systems%2C+Clinical%22%5BMesh%5D&sort=pubdate&size=20&ac=no) |
| #20 | Search: **"Decision Making"[Mesh:NoExp]** Sort by: **Publication Date** | [103,752](https://pubmed.ncbi.nlm.nih.gov/?term=%22Decision+Making%22%5BMesh%3ANoExp%5D&sort=pubdate&size=20&ac=no) |
| #19 | Search: **"Decision Support Techniques"[Mesh:NoExp]** Sort by: **Publication Date** | [22,364](https://pubmed.ncbi.nlm.nih.gov/?term=%22Decision+Support+Techniques%22%5BMesh%3ANoExp%5D&sort=pubdate&size=20&ac=no) |
| #18 | Search: **"Decision Making, Shared"[Mesh]** Sort by: **Publication Date** | [1,783](https://pubmed.ncbi.nlm.nih.gov/?term=%22Decision+Making%2C+Shared%22%5BMesh%5D&sort=pubdate&size=20&ac=no) |
| #17 | Search: **(((((((((((("Musculoskeletal Pain"[Mesh]) OR ("Musculoskeletal Diseases"[Mesh])) OR ("Arthralgia"[Mesh])) OR ("Back Pain"[Mesh])) OR ("Facial Pain"[Mesh])) OR ("Headache"[Mesh])) OR ("Metatarsalgia"[Mesh])) OR ("Neck Pain"[Mesh])) OR ("Nociceptive Pain"[Mesh])) OR ("Pain, Intractable"[Mesh])) OR ("Pelvic Pain"[Mesh])) OR (musculoskeletal[Text Word] OR hip pain*[Text Word] OR knee pain*[Text Word] OR shoulder pain*[Text Word] OR neck pain*[Text Word] OR back pain*[Text Word] OR elbow pain*[Text Word] OR hand pain*[Text Word] OR headach*[Text Word] OR pelvic pain*[Text Word] OR facial pain*[Text Word] OR leg pain*[Text Word] OR bursitis[Text Word] OR tendin*[Text Word] OR tendon*[Text Word] OR myalgia[Text Word] OR arthralgia*[Text Word] OR joint pain*[Text Word] OR bone pain*[Text Word] OR ostalgia[Text Word] OR "Fractures, Bone"[Mesh] OR fracture*[Text Word] OR arthrit*[Text Word] OR rheumat*[Text Word] OR arthro*[Text Word] OR arm pain*[Text Word] OR muscle pain*[Text Word] OR "Soft Tissue Injuries"[Mesh] OR foot pain*[Text Word])) AND (("Chronic Pain"[Mesh]) OR ((chronic*[Text Word] OR recurr*[Text Word] OR continu*[Text Word] OR longstanding[Text Word] OR persist*[Text Word]) AND pain*[Text Word]))** Sort by: **Publication Date** | [105,489](https://pubmed.ncbi.nlm.nih.gov/?term=%28%28%28%28%28%28%28%28%28%28%28%28%22Musculoskeletal+Pain%22%5BMesh%5D%29+OR+%28%22Musculoskeletal+Diseases%22%5BMesh%5D%29%29+OR+%28%22Arthralgia%22%5BMesh%5D%29%29+OR+%28%22Back+Pain%22%5BMesh%5D%29%29+OR+%28%22Facial+Pain%22%5BMesh%5D%29%29+OR+%28%22Headache%22%5BMesh%5D%29%29+OR+%28%22Metatarsalgia%22%5BMesh%5D%29%29+OR+%28%22Neck+Pain%22%5BMesh%5D%29%29+OR+%28%22Nociceptive+Pain%22%5BMesh%5D%29%29+OR+%28%22Pain%2C+Intractable%22%5BMesh%5D%29%29+OR+%28%22Pelvic+Pain%22%5BMesh%5D%29%29+OR+%28musculoskeletal%5BText+Word%5D+OR+hip+pain%2A%5BText+Word%5D+OR+knee+pain%2A%5BText+Word%5D+OR+shoulder+pain%2A%5BText+Word%5D+OR+neck+pain%2A%5BText+Word%5D+OR+back+pain%2A%5BText+Word%5D+OR+elbow+pain%2A%5BText+Word%5D+OR+hand+pain%2A%5BText+Word%5D+OR+headach%2A%5BText+Word%5D+OR+pelvic+pain%2A%5BText+Word%5D+OR+facial+pain%2A%5BText+Word%5D+OR+leg+pain%2A%5BText+Word%5D+OR+bursitis%5BText+Word%5D+OR+tendin%2A%5BText+Word%5D+OR+tendon%2A%5BText+Word%5D+OR+myalgia%5BText+Word%5D+OR+arthralgia%2A%5BText+Word%5D+OR+joint+pain%2A%5BText+Word%5D+OR+bone+pain%2A%5BText+Word%5D+OR+ostalgia%5BText+Word%5D+OR+%22Fractures%2C+Bone%22%5BMesh%5D+OR+fracture%2A%5BText+Word%5D+OR+arthrit%2A%5BText+Word%5D+OR+rheumat%2A%5BText+Word%5D+OR+arthro%2A%5BText+Word%5D+OR+arm+pain%2A%5BText+Word%5D+OR+muscle+pain%2A%5BText+Word%5D+OR+%22Soft+Tissue+Injuries%22%5BMesh%5D+OR+foot+pain%2A%5BText+Word%5D%29%29+AND+%28%28%22Chronic+Pain%22%5BMesh%5D%29+OR+%28%28chronic%2A%5BText+Word%5D+OR+recurr%2A%5BText+Word%5D+OR+continu%2A%5BText+Word%5D+OR+longstanding%5BText+Word%5D+OR+persist%2A%5BText+Word%5D%29+AND+pain%2A%5BText+Word%5D%29%29&sort=pubdate&size=20&ac=no) |
| #16 | Search: **("Chronic Pain"[Mesh]) OR ((chronic*[Text Word] OR recurr*[Text Word] OR continu*[Text Word] OR longstanding[Text Word] OR persist*[Text Word]) AND pain*[Text Word])** Sort by: **Publication Date** | [261,961](https://pubmed.ncbi.nlm.nih.gov/?term=%28%22Chronic+Pain%22%5BMesh%5D%29+OR+%28%28chronic%2A%5BText+Word%5D+OR+recurr%2A%5BText+Word%5D+OR+continu%2A%5BText+Word%5D+OR+longstanding%5BText+Word%5D+OR+persist%2A%5BText+Word%5D%29+AND+pain%2A%5BText+Word%5D%29&sort=pubdate&size=20&ac=no) |
| #15 | Search: **(chronic*[Text Word] OR recurr*[Text Word] OR continu*[Text Word] OR longstanding[Text Word] OR persist*[Text Word]) AND pain*[Text Word]** Sort by: **Publication Date** | [261,961](https://pubmed.ncbi.nlm.nih.gov/?term=%28chronic%2A%5BText+Word%5D+OR+recurr%2A%5BText+Word%5D+OR+continu%2A%5BText+Word%5D+OR+longstanding%5BText+Word%5D+OR+persist%2A%5BText+Word%5D%29+AND+pain%2A%5BText+Word%5D&sort=pubdate&size=20&ac=no) |
| #14 | Search: **"Chronic Pain"[Mesh]** Sort by: **Publication Date** | [21,297](https://pubmed.ncbi.nlm.nih.gov/?term=%22Chronic+Pain%22%5BMesh%5D&sort=pubdate&size=20&ac=no) |
| #13 | Search: **((((((((((("Musculoskeletal Pain"[Mesh]) OR ("Musculoskeletal Diseases"[Mesh])) OR ("Arthralgia"[Mesh])) OR ("Back Pain"[Mesh])) OR ("Facial Pain"[Mesh])) OR ("Headache"[Mesh])) OR ("Metatarsalgia"[Mesh])) OR ("Neck Pain"[Mesh])) OR ("Nociceptive Pain"[Mesh])) OR ("Pain, Intractable"[Mesh])) OR ("Pelvic Pain"[Mesh])) OR (musculoskeletal[Text Word] OR hip pain*[Text Word] OR knee pain*[Text Word] OR shoulder pain*[Text Word] OR neck pain*[Text Word] OR back pain*[Text Word] OR elbow pain*[Text Word] OR hand pain*[Text Word] OR headach*[Text Word] OR pelvic pain*[Text Word] OR facial pain*[Text Word] OR leg pain*[Text Word] OR bursitis[Text Word] OR tendin*[Text Word] OR tendon*[Text Word] OR myalgia[Text Word] OR arthralgia*[Text Word] OR joint pain*[Text Word] OR bone pain*[Text Word] OR ostalgia[Text Word] OR "Fractures, Bone"[Mesh] OR fracture*[Text Word] OR arthrit*[Text Word] OR rheumat*[Text Word] OR arthro*[Text Word] OR arm pain*[Text Word] OR muscle pain*[Text Word] OR "Soft Tissue Injuries"[Mesh] OR foot pain*[Text Word])** Sort by: **Publication Date** | [1,982,063](https://pubmed.ncbi.nlm.nih.gov/?term=%28%28%28%28%28%28%28%28%28%28%28%22Musculoskeletal+Pain%22%5BMesh%5D%29+OR+%28%22Musculoskeletal+Diseases%22%5BMesh%5D%29%29+OR+%28%22Arthralgia%22%5BMesh%5D%29%29+OR+%28%22Back+Pain%22%5BMesh%5D%29%29+OR+%28%22Facial+Pain%22%5BMesh%5D%29%29+OR+%28%22Headache%22%5BMesh%5D%29%29+OR+%28%22Metatarsalgia%22%5BMesh%5D%29%29+OR+%28%22Neck+Pain%22%5BMesh%5D%29%29+OR+%28%22Nociceptive+Pain%22%5BMesh%5D%29%29+OR+%28%22Pain%2C+Intractable%22%5BMesh%5D%29%29+OR+%28%22Pelvic+Pain%22%5BMesh%5D%29%29+OR+%28musculoskeletal%5BText+Word%5D+OR+hip+pain%2A%5BText+Word%5D+OR+knee+pain%2A%5BText+Word%5D+OR+shoulder+pain%2A%5BText+Word%5D+OR+neck+pain%2A%5BText+Word%5D+OR+back+pain%2A%5BText+Word%5D+OR+elbow+pain%2A%5BText+Word%5D+OR+hand+pain%2A%5BText+Word%5D+OR+headach%2A%5BText+Word%5D+OR+pelvic+pain%2A%5BText+Word%5D+OR+facial+pain%2A%5BText+Word%5D+OR+leg+pain%2A%5BText+Word%5D+OR+bursitis%5BText+Word%5D+OR+tendin%2A%5BText+Word%5D+OR+tendon%2A%5BText+Word%5D+OR+myalgia%5BText+Word%5D+OR+arthralgia%2A%5BText+Word%5D+OR+joint+pain%2A%5BText+Word%5D+OR+bone+pain%2A%5BText+Word%5D+OR+ostalgia%5BText+Word%5D+OR+%22Fractures%2C+Bone%22%5BMesh%5D+OR+fracture%2A%5BText+Word%5D+OR+arthrit%2A%5BText+Word%5D+OR+rheumat%2A%5BText+Word%5D+OR+arthro%2A%5BText+Word%5D+OR+arm+pain%2A%5BText+Word%5D+OR+muscle+pain%2A%5BText+Word%5D+OR+%22Soft+Tissue+Injuries%22%5BMesh%5D+OR+foot+pain%2A%5BText+Word%5D%29&sort=pubdate&size=20&ac=no) |
| #12 | Search: **musculoskeletal[Text Word] OR hip pain*[Text Word] OR knee pain*[Text Word] OR shoulder pain*[Text Word] OR neck pain*[Text Word] OR back pain*[Text Word] OR elbow pain*[Text Word] OR hand pain*[Text Word] OR headach*[Text Word] OR pelvic pain*[Text Word] OR facial pain*[Text Word] OR leg pain*[Text Word] OR bursitis[Text Word] OR tendin*[Text Word] OR tendon*[Text Word] OR myalgia[Text Word] OR arthralgia*[Text Word] OR joint pain*[Text Word] OR bone pain*[Text Word] OR ostalgia[Text Word] OR "Fractures, Bone"[Mesh] OR fracture*[Text Word] OR arthrit*[Text Word] OR rheumat*[Text Word] OR arthro*[Text Word] OR arm pain*[Text Word] OR muscle pain*[Text Word] OR "Soft Tissue Injuries"[Mesh] OR foot pain*[Text Word]** Sort by: **Publication Date** | [1,237,552](https://pubmed.ncbi.nlm.nih.gov/?term=musculoskeletal%5BText+Word%5D+OR+hip+pain%2A%5BText+Word%5D+OR+knee+pain%2A%5BText+Word%5D+OR+shoulder+pain%2A%5BText+Word%5D+OR+neck+pain%2A%5BText+Word%5D+OR+back+pain%2A%5BText+Word%5D+OR+elbow+pain%2A%5BText+Word%5D+OR+hand+pain%2A%5BText+Word%5D+OR+headach%2A%5BText+Word%5D+OR+pelvic+pain%2A%5BText+Word%5D+OR+facial+pain%2A%5BText+Word%5D+OR+leg+pain%2A%5BText+Word%5D+OR+bursitis%5BText+Word%5D+OR+tendin%2A%5BText+Word%5D+OR+tendon%2A%5BText+Word%5D+OR+myalgia%5BText+Word%5D+OR+arthralgia%2A%5BText+Word%5D+OR+joint+pain%2A%5BText+Word%5D+OR+bone+pain%2A%5BText+Word%5D+OR+ostalgia%5BText+Word%5D+OR+%22Fractures%2C+Bone%22%5BMesh%5D+OR+fracture%2A%5BText+Word%5D+OR+arthrit%2A%5BText+Word%5D+OR+rheumat%2A%5BText+Word%5D+OR+arthro%2A%5BText+Word%5D+OR+arm+pain%2A%5BText+Word%5D+OR+muscle+pain%2A%5BText+Word%5D+OR+%22Soft+Tissue+Injuries%22%5BMesh%5D+OR+foot+pain%2A%5BText+Word%5D&sort=pubdate&size=20&ac=no) |
| #11 | Search: **"Pelvic Pain"[Mesh]** Sort by: **Publication Date** | [10,864](https://pubmed.ncbi.nlm.nih.gov/?term=%22Pelvic+Pain%22%5BMesh%5D&sort=pubdate&size=20&ac=no) |
| #10 | Search: **"Pain, Intractable"[Mesh]** Sort by: **Publication Date** | [6,356](https://pubmed.ncbi.nlm.nih.gov/?term=%22Pain%2C+Intractable%22%5BMesh%5D&sort=pubdate&size=20&ac=no) |
| #9 | Search: **"Nociceptive Pain"[Mesh]** Sort by: **Publication Date** | [1,710](https://pubmed.ncbi.nlm.nih.gov/?term=%22Nociceptive+Pain%22%5BMesh%5D&sort=pubdate&size=20&ac=no) |
| #8 | Search: **"Neck Pain"[Mesh]** Sort by: **Publication Date** | [8,299](https://pubmed.ncbi.nlm.nih.gov/?term=%22Neck+Pain%22%5BMesh%5D&sort=pubdate&size=20&ac=no) |
| #7 | Search: **"Metatarsalgia"[Mesh]** Sort by: **Publication Date** | [422](https://pubmed.ncbi.nlm.nih.gov/?term=%22Metatarsalgia%22%5BMesh%5D&sort=pubdate&size=20&ac=no) |
| #6 | Search: **"Headache"[Mesh]** Sort by: **Publication Date** | [30,876](https://pubmed.ncbi.nlm.nih.gov/?term=%22Headache%22%5BMesh%5D&sort=pubdate&size=20&ac=no) |
| #5 | Search: **"Facial Pain"[Mesh]** Sort by: **Publication Date** | [9,698](https://pubmed.ncbi.nlm.nih.gov/?term=%22Facial+Pain%22%5BMesh%5D&sort=pubdate&size=20&ac=no) |
| #4 | Search: **"Back Pain"[Mesh]** Sort by: **Publication Date** | [44,126](https://pubmed.ncbi.nlm.nih.gov/?term=%22Back+Pain%22%5BMesh%5D&sort=pubdate&size=20&ac=no) |
| #3 | Search: **"Arthralgia"[Mesh]** Sort by: **Publication Date** | [15,359](https://pubmed.ncbi.nlm.nih.gov/?term=%22Arthralgia%22%5BMesh%5D&sort=pubdate&size=20&ac=no) |
| #2 | Search: **"Musculoskeletal Diseases"[Mesh]** Sort by: **Publication Date** | [1,192,324](https://pubmed.ncbi.nlm.nih.gov/?term=%22Musculoskeletal+Diseases%22%5BMesh%5D&sort=pubdate&size=20&ac=no) |
| #1 | Search: **"Musculoskeletal Pain"[Mesh]** Sort by: **Publication Date** | [7,150](https://pubmed.ncbi.nlm.nih.gov/?term=%22Musculoskeletal+Pain%22%5BMesh%5D&sort=pubdate&size=20&ac=no) |

### Embase

| No. | Query | Results |
| --- | --- | --- |
| #36 | #25 AND #35 | 1499 |
| #35 | #26 OR #27 OR #28 OR #29 OR #30 OR #31 OR #32 OR #33 OR #34 | 516633 |
| #34 | ((patient* OR consumer*) NEAR/4 (participat* OR involv* OR interaction*)):ti | 13864 |
| #33 | 'consumer participation*':ti,ab,kw OR 'consumer involvement*':ti,ab,kw | 1141 |
| #32 | 'patient participation*':ti,ab,kw OR 'patient involvement*':ti,ab,kw | 9261 |
| #31 | 'patient participation'/de | 33986 |
| #30 | ((decision OR choice) NEAR/2 (making* OR support* OR aid* OR behavio*)):ti,ab,kw | 288681 |
| #29 | ((share* OR sharing* OR informed*) NEAR/3 (decision* OR deciding* OR choice*)):ti,ab,kw | 43600 |
| #28 | 'decision making'/de | 271089 |
| #27 | 'decision support system'/exp | 32548 |
| #26 | 'shared decision making'/de | 12982 |
| #25 | #23 OR #24 | 93282 |
| #24 | 'chronic musculoskeletal pain'/de | 36 |
| #23 | #19 AND #22 | 93282 |
| #22 | #20 OR #21 | 183258 |
| #21 | ((chronic* OR recurr* OR continu* OR longstanding OR persist*) NEAR/4 pain*):ti,ab,kw | 166903 |
| #20 | 'chronic pain'/de | 73719 |
| #19 | #1 OR #2 OR #3 OR #4 OR #5 OR #6 OR #7 OR #8 OR #9 OR #10 OR #11 OR #12 OR #13 OR #14 OR #15 OR #16 OR #17 OR #18 | 3440121 |
| #18 | bursitis:ti,ab,kw | 4333 |
| #17 | headach*:ti,ab,kw | 163141 |
| #16 | fracture*:ti,ab,kw | 374824 |
| #15 | arthrit*:ti,ab,kw OR rheumat*:ti,ab,kw OR arthro*:ti,ab,kw | 636201 |
| #14 | arthralgia*:ti,ab,kw OR ostalgia:ti,ab,kw | 18984 |
| #13 | tendin*:ti,ab,kw OR tendon*:ti,ab,kw | 108862 |
| #12 | myalgia:ti,ab,kw | 14710 |
| #11 | ((hip OR knee OR shoulder OR neck OR back OR elbow OR hand OR pelvic OR facial OR leg OR foot OR bone OR arm OR muscle OR joint OR musculoskeletal) NEAR/1 pain*):ti,ab,kw | 209034 |
| #10 | 'soft tissue injury'/exp | 11036 |
| #9 | 'fracture'/exp | 379229 |
| #8 | 'myalgia'/exp | 129694 |
| #7 | 'pelvic pain'/exp | 23391 |
| #6 | 'intractable pain'/de | 5455 |
| #5 | 'nociceptive pain'/de | 2185 |
| #4 | 'metatarsalgia'/de | 1380 |
| #3 | 'headache and facial pain'/exp | 370827 |
| #2 | 'musculoskeletal disease'/exp | 2808728 |
| #1 | 'musculoskeletal pain'/exp | 183279 |

### Cochrane Library

| ID | Search | Hits |
| --- | --- | --- |
| #1 | MeSH descriptor: [Musculoskeletal Pain] explode all trees | 1233 |
| #2 | MeSH descriptor: [Arthralgia] explode all trees | 2165 |
| #3 | MeSH descriptor: [Back Pain] explode all trees | 5855 |
| #4 | MeSH descriptor: [Facial Pain] explode all trees | 818 |
| #5 | MeSH descriptor: [Headache] explode all trees | 2649 |
| #6 | MeSH descriptor: [Metatarsalgia] explode all trees | 22 |
| #7 | MeSH descriptor: [Neck Pain] explode all trees | 1604 |
| #8 | MeSH descriptor: [Nociceptive Pain] explode all trees | 130 |
| #9 | MeSH descriptor: [Pain, Intractable] explode all trees | 278 |
| #10 | MeSH descriptor: [Pelvic Pain] explode all trees | 1383 |
| #11 | MeSH descriptor: [Musculoskeletal Diseases] explode all trees | 46639 |
| #12 | MeSH descriptor: [Myalgia] explode all trees | 616 |
| #13 | MeSH descriptor: [Fractures, Bone] explode all trees | 6989 |
| #14 | MeSH descriptor: [Soft Tissue Injuries] explode all trees | 144 |
| #15 | MeSH descriptor: [Bursitis] explode all trees | 482 |
| #16 | ((hip OR knee OR shoulder OR neck OR back OR elbow OR hand OR pelvic OR facial OR leg OR foot OR bone OR arm OR muscle OR joint OR musculoskeletal) NEAR/1 pain):ti,ab,kw | 40749 |
| #17 | (tendin* OR tendon*):ti,ab,kw | 7235 |
| #18 | (myalgia OR arthralgia* OR ostalgia):ti,ab,kw | 9748 |
| #19 | (arthrit* OR rheumat* OR arthro*):ti,ab,kw | 52689 |
| #20 | (bursitis OR fracture*):ti,ab,kw | 27569 |
| #21 | (headach*):ti,ab,kw | 35830 |
| #22 | {OR #1-#21} | 175053 |
| #23 | MeSH descriptor: [Chronic Pain] explode all trees | 3235 |
| #24 | ((chronic* OR recurr* OR continu* OR longstanding OR persist*) NEAR/4 pain):ti,ab,kw | 25423 |
| #25 | {OR #23-#24} | 25423 |
| #26 | #22 AND #25 | 14461 |
| #27 | MeSH descriptor: [Decision Making, Shared] explode all trees | 92 |
| #28 | MeSH descriptor: [Decision Support Techniques] this term only | 912 |
| #29 | MeSH descriptor: [Decision Making] this term only | 2346 |
| #30 | MeSH descriptor: [Decision Support Systems, Clinical] explode all trees | 458 |
| #31 | MeSH descriptor: [Choice Behavior] this term only | 1390 |
| #32 | ((shared or sharing) NEXT decision*):ti,ab,kw | 1908 |
| #33 | (informed NEXT (choice* or decision*)):ti,ab,kw | 1354 |
| #34 | (decision NEXT aid*):ti,ab,kw | 1637 |
| #35 | ((share* or sharing* or informed*) NEAR/3 (decision* or deciding* or choice*)):ti,ab,kw | 3694 |
| #36 | ((decision OR choice) NEAR/2 (making* OR support* OR aid* OR behavio*)):ti,ab,kw | 21159 |
| #37 | MeSH descriptor: [Patient Participation] explode all trees | 1541 |
| #38 | ((patient or consumer) NEXT (participat* or involv*)):ti,ab,kw | 3828 |
| #39 | ((patient* or consumer*) NEAR/4 (participat* or involv* or interaction*)):ti | 1116 |
| #40 | {OR #27-#39} | 25797 |
| #41 | #26 AND #40 | 231 |

### Scopus

| # | Query | Hits |
| --- | --- | --- |
| #3 | #1 AND #2 | 972 |
| #2 | ( TITLE-ABS-KEY ( "shared decision*" OR "sharing decision*" ) OR TITLE-ABS-KEY ( "informed decision*" OR "informed choice*" OR "decision aid*" ) OR TITLE-ABS-KEY ( ( ( share* OR sharing* OR informed* ) AND ( decision* OR deciding* OR choice* ) ) ) OR TITLE-ABS-KEY ( "decision making*" OR "decision support*" OR "choice behavio*" ) OR TITLE ( ( ( decision* OR choice* ) AND ( making* OR support* OR behavio* ) ) ) OR TITLE-ABS-KEY ( "patient participation*" OR "consumer participation*" OR "patient involvement*" OR "consumer involvement*" ) OR TITLE ( ( ( patient* OR consumer* ) AND ( involvement* OR involving* OR participation* OR participating* OR interact* ) ) ) ) | 33,500 |
| #1 | ( TITLE-ABS-KEY ( ( "musculoskeletal pain" OR "hip pain*" OR "knee pain*" OR "shoulder pain*" OR "neck pain*" OR "back pain*" OR "elbow pain*" OR "hand pain*" OR headach* OR "pelvic pain*" OR "facial pain*" OR "leg pain*" OR bursitis OR tendin* OR tendon* OR myalgia OR arthralgia* OR "joint pain*" OR "bone pain*" OR ostalgia OR fracture* OR arthrit* OR rheumat* OR arthro* OR "arm pain*" OR "muscle pain*" OR "soft tissue injuries" OR "foot pain*" ) ) AND TITLE-ABS-KEY ( "chronic pain*" OR "recurrent pain*" OR "continuing pain*" OR "continuous pain*" OR "continuously pain*" OR "longstanding pain*" OR "persisting pain*" ) ) | 1,276,739 |

### PsycINFO

| # | Query | Hits |
| --- | --- | --- |
| #3 | #1 AND #2 | 23 |
| #2 | ((IndexTermsFilt: ("Decision Making") OR IndexTermsFilt: ("Choice Behavior") OR IndexTermsFilt: ("Group Decision Making") OR IndexTermsFilt: ("Management Decision Making") OR IndexTermsFilt: ("Decision Support Systems"))) OR ((title: (share*) OR title: (sharing*) OR title: (informed*)) NEAR/3 (title: (decision*) OR title: (deciding*) OR title: (choice*)) OR (abstract: (share*) OR abstract: (sharing*) OR abstract: (informed*)) NEAR/3 (abstract: (decision*) OR abstract: (deciding*) OR abstract: (choice*)) OR (Keywords: (share*) OR Keywords: (sharing*) OR Keywords: (informed*)) NEAR/3 (Keywords: (decision*) OR Keywords: (deciding*) OR Keywords: (choice*))) OR ((title: (decision) OR title: (choice)) NEAR/2 (title: (making*) OR title: (support*) OR title: (aid*) OR title: (behavio*)) OR (abstract: (decision) OR abstract: (choice)) NEAR/2 (abstract: (making*) OR abstract: (support*) OR abstract: (aid*) OR abstract: (behavio*)) OR (Keywords: (decision) OR Keywords: (choice)) NEAR/2 (Keywords: (making*) OR Keywords: (support*) OR Keywords: (aid*) OR Keywords: (behavio*))) OR ((IndexTermsFilt: ("Client Participation"))) OR ((title: ("patient participat*") OR title: ("consumer participat*") OR title: ("patient involv*") OR title: ("consumer involv*")) OR (abstract: ("patient participat*") OR abstract: ("consumer participat*") OR abstract: ("patient involv*") OR abstract: ("consumer involv*")) OR (Keywords: ("patient participat*") OR Keywords: ("consumer participat*") OR Keywords: ("patient involv*") OR Keywords: ("consumer involv*"))) OR ((title: (patient*) OR title: (consumer*)) NEAR/4 (title: (participat*) OR title: (involv*) OR title: (interaction*))) | 171313 |
| #1 | ((((((IndexTermsFilt: ("Chronic Pain"))))) OR ((((title: (chronic*))) OR ((title: (recurr*))) OR ((title: (continu*))) OR ((title: (longstanding))) OR ((title: (persist*)))) NEAR/4 ((title: (pain))) OR (((abstract: (chronic*))) OR ((abstract: (recurr*))) OR ((abstract: (continu*))) OR ((abstract: (longstanding))) OR ((abstract: (persist*)))) NEAR/4 ((abstract: (pain))) OR (((Keywords: (chronic*))) OR ((Keywords: (recurr*))) OR ((Keywords: (continu*))) OR ((Keywords: (longstanding))) OR ((Keywords: (persist*)))) NEAR/4 ((Keywords: (pain))))) AND (((((IndexTermsFilt: ("Pain"))) OR ((IndexTermsFilt: ("Back Pain"))) OR ((IndexTermsFilt: ("Headache"))) OR ((IndexTermsFilt: ("Fibromyalgia"))) OR ((IndexTermsFilt: ("Muscular Disorders"))))) OR ((((title: (hip))) OR ((title: (knee))) OR ((title: (shoulder))) OR ((title: (neck))) OR ((title: (back))) OR ((title: (elbow))) OR ((title: (hand))) OR ((title: (pelvic))) OR ((title: (facial))) OR ((title: (leg))) OR ((title: (foot))) OR ((title: (bone))) OR ((title: (arm))) OR ((title: (muscle))) OR ((title: (joint))) OR ((title: (musculoskeletal)))) NEAR/1 ((title: (pain))) OR (((abstract: (hip))) OR ((abstract: (knee))) OR ((abstract: (shoulder))) OR ((abstract: (neck))) OR ((abstract: (back))) OR ((abstract: (elbow))) OR ((abstract: (hand))) OR ((abstract: (pelvic))) OR ((abstract: (facial))) OR ((abstract: (leg))) OR ((abstract: (foot))) OR ((abstract: (bone))) OR ((abstract: (arm))) OR ((abstract: (muscle))) OR ((abstract: (joint))) OR ((abstract: (musculoskeletal)))) NEAR/1 ((abstract: (pain))) OR (((Keywords: (hip))) OR ((Keywords: (knee))) OR ((Keywords: (shoulder))) OR ((Keywords: (neck))) OR ((Keywords: (back))) OR ((Keywords: (elbow))) OR ((Keywords: (hand))) OR ((Keywords: (pelvic))) OR ((Keywords: (facial))) OR ((Keywords: (leg))) OR ((Keywords: (foot))) OR ((Keywords: (bone))) OR ((Keywords: (arm))) OR ((Keywords: (muscle))) OR ((Keywords: (joint))) OR ((Keywords: (musculoskeletal)))) NEAR/1 ((Keywords: (pain)))) OR ((((title: (tendin*))) OR ((title: (tendon*))) OR ((title: (myalgia))) OR ((title: (arthralgia*))) OR ((title: (ostalgia))) OR ((title: (arthrit*))) OR ((title: (rheumat*))) OR ((title: (arthro*))) OR ((title: (bursitis))) OR ((title: (fracture*))) OR ((title: (headach*)))) OR (((abstract: (tendin*))) OR ((abstract: (tendon*))) OR ((abstract: (myalgia))) OR ((abstract: (arthralgia*))) OR ((abstract: (ostalgia))) OR ((abstract: (arthrit*))) OR ((abstract: (rheumat*))) OR ((abstract: (arthro*))) OR ((abstract: (bursitis))) OR ((abstract: (fracture*))) OR ((abstract: (headach*)))) OR (((Keywords: (tendin*))) OR ((Keywords: (tendon*))) OR ((Keywords: (myalgia))) OR ((Keywords: (arthralgia*))) OR ((Keywords: (ostalgia))) OR ((Keywords: (arthrit*))) OR ((Keywords: (rheumat*))) OR ((Keywords: (arthro*))) OR ((Keywords: (bursitis))) OR ((Keywords: (fracture*))) OR ((Keywords: (headach*))))))) AND ((((IndexTermsFilt:("Systematic Review")) OR (IndexTermsFilt:("Meta Analysis")))) OR (((title:("meta analy*")) OR (title:("meta-analy*")) OR (title:(metaanaly*))) OR ((abstract:("meta analy*")) OR (abstract:("meta-analy*")) OR (abstract:(metaanaly*))) OR ((Subject:("meta analy*")) OR (Subject:("meta-analy*")) OR (Subject:(metaanaly*)))) OR (((Title:(systematic NEAR/3) (Title:(review*) OR Title:(overview*) OR Title:(study) OR Title:(studies) OR Title:(search*) OR Title:(approach*))) OR (Abstract:(systematic NEAR/3) (Abstract:(review*) OR Abstract:(overview*) OR Abstract:(study) OR Abstract:(studies) OR Abstract:(search*) OR Abstract:(approach*))) OR (Subject:(systematic NEAR/3) (Subject:(review*) OR Subject:(overview*) OR Subject:(study) OR Subject:(studies) OR Subject:(search*) OR Subject:(approach*)))))) | 718 |

### CINAHL with Fulltext

| # | Query | Results |
| --- | --- | --- |
| S24 | S16 AND S23 | 654 |
| S23 | S17 OR S18 OR S19 OR S20 OR S21 OR S22 | 225,579 |
| S22 | TI (patient* OR consumer*) N4 (participat* OR involv* OR interaction*) | 6,000 |
| S21 | ( patient participation* OR patient involvement* ) OR ( consumer participation* OR consumer involvement* ) | 26,374 |
| S20 | (MH "Consumer Participation") | 23,685 |
| S19 | (decision OR choice) N2 (making* OR support* OR aid* OR behavio*) | 189,017 |
| S18 | (share* OR sharing* OR informed*) N3 (decision* OR deciding* OR choice*) | 18,605 |
| S17 | (MH "Decision Making, Shared") OR (MH "Decision Support Techniques+") OR (MH "Decision Making, Patient+") OR (MH "Decision Making") | 91,910 |
| S16 | S12 AND S15 | 28,379 |
| S15 | S13 OR S14 | 58,113 |
| S14 | (chronic* OR recurr* OR continu* OR longstanding OR persist*) N4 pain | 58,113 |
| S13 | (MH "Chronic Pain") | 26,233 |
| S12 | S1 OR S2 OR S3 OR S4 OR S5 OR S6 OR S7 OR S8 OR S9 OR S10 OR S11 | 571,195 |
| S11 | headach* | 36,826 |
| S10 | bursitis OR fracture* | 102,809 |
| S9 | arthrit* OR rheumat* OR arthro* | 174,343 |
| S8 | myalgia OR arthralgia* OR ostalgia | 7,305 |
| S7 | tendin* OR tendon* | 28,395 |
| S6 | (hip OR knee OR shoulder OR neck OR back OR elbow OR hand OR pelvic OR facial OR leg OR foot OR bone OR arm OR muscle OR joint OR musculoskeletal) N1 pain | 91,324 |
| S5 | (MH "Soft Tissue Injuries+") | 2,740 |
| S4 | (MH "Fractures+") | 67,950 |
| S3 | (MH "Musculoskeletal Diseases+") | 307,970 |
| S2 | (MH "Bursitis+") | 2,005 |
| S1 | (MH "Back Pain+") OR (MH "Facial Pain") OR (MH "Headache") OR (MH "Knee Pain+") OR (MH "Metatarsalgia") OR (MH "Muscle Pain") OR (MH "Neck Pain") OR (MH "Nociceptive Pain+") OR (MH "Arthralgia+") OR (MH "Elbow Pain") OR (MH "Heel Pain") OR (MH "Shoulder Pain") OR (MH "Pelvic Pain+") | 77,439 |

### PEDro

| Search Fields | Search | Results |
| --- | --- | --- |
| Abstract & Title | Shared decision making (16)  Patient participation (25)  Informed decision (7)  Decision support (20)  Patient involvement (11)  Patient interaction (46)  Informed choice (2) | Found 109 records |
| Subdiscipline | Musculoskeletal |  |
| Topic | Chronic pain |  |
