## Appendix 2 for "Implementation of Shared Decision-Making in the Management of Chronic Musculoskeletal Pain: *a scoping review*"

| **#** | **Author/ date** | **Journal** | **Title** | **Study objective** | **Design** | **Data source** | **Participants** |
| --- | --- | --- | --- | --- | --- | --- | --- |
| **1** | Alotaibi, 2023, Saudi Arabia | Journal of Multidisciplinary Healthcare | The Practice of Shared Decision-Making Among Physiotherapists and Patients with Musculoskeletal Conditions | This study aimed to explore how SDM between patients and physiotherapists may lead to patient satisfaction. | Cross-sectional study | Questionnaire | 106 patients with MSK pain and 9 physiotherapists. |
| **2** | Arnborg Lund, 2020, Denmark | Journal of Rehabilitation Medicine | Communicating and diagnosing non-specific low back pain: A qualitative study of the healthcare | To explore, from the perspective of healthcare practitioners, how persistent non- specific low back pain may be communicated in a way that moves beyond a biomedical diagnosis | Qualitative | Semi-structured focus group interviews and individual interviews. | 10 HCP (physiotherapists, chiropractors and general practitioners). |
| **2** | Bieber, 2006, Germany | Patient Education and Counseling | Long-term effects of a shared decision-making intervention on physician-patient interaction and outcome in fibromyalgia. A qualitative and quantitative 1 year follow-up of a randomized controlled trial | To investigate the effects of a shared decision-making (SDM) intervention on physician–patient interaction and health outcome. | Mixed method follow up of randomized controlled trial | Qualitative:  Doctors: structured protocol after each patient contact Patients: semi-standardized interviews  Quantitative:  Patients: FAPI questionnaire  Doctors: DDPRQ questionnaire | 111 patients with FMS  (34 SDM group, 33 information group and 44 comparison group).  13 HCP (doctors)  (3 treating SDM group, 6 treating information group and 3 treating comparison group). |
| **3** | Bieber, 2008, Germany | Journal of Psychosomathic Research | A shared decision-making communication training program for physicians treating fibromyalgia patients: effects of a randomized controlled trial | To assess whether SDM improves the quality of physician– patient interaction from patients’ perspective. | Randomised controlled trial | Questionnaires | 85 patients with fibromyalgia (44 in SDM group, 41 in information-only group). |
| **44** | Boyle, 2022, Australia | Musculoskeletal Science and Practice | Patient perspectives of care pathways for people with low back pain: A qualitative study | To explore patients’ perspectives and/or experience of care pathways and their involvement in decision- making in primary care. | Qualitative | Semi-structured interviews | 14 patients with LBP. |
| **55** | Camerini, 2016, Switzerland | Patient Education and Counseling | Patients' need for information provision and perceived participation in decision making in doctor-patient consultation: Micro-cultural differences between French- and Italian-speaking Switzerland | To explore micro-cultural differences in patients’ need for information provision, perceived participation in decision making, and related concepts during the doctor-patient consultation between French- and Italian-speaking patients in Switzerland. | Cross-sectional survey | Questionnaire | 237 patients with cLBP. |
| **66** | Chen, 2021, Taiwan | Patient Education and Counseling | Effectiveness of shared decision-making intervention in patients with lumbar degenerative diseases: a randomized controlled trial [with consumer summary] | To evaluate the efficacy of shared decision-making (SDM) intervention among patients with lumbar degenerative diseases (LDDs) in terms of decision self-efficacy, control preferences, SDM process, decision satisfaction, and conflict. | Randomised controlled trial | Questionnaires | 130 patients (67 SDM intervention, 63 comparison group). |
| **87** | Cooper, 2008, Scotland | Physiotherapy | Patient-centredness in physiotherapy from the perspective of the chronic low back pain patient | To define patient-centredness from the patient’s perspective in the context of physiotherapy for chronic low back pain (CLBP). | Qualitative | Semi-structured interviews | 25 patients with CLBP. |
| **08** | Coutu, 2015, Canada | Patient Education and Counseling | Occupational therapists' shared decision-making behaviors with patients having persistent pain in a work rehabilitation context: A cross-sectional study. | To asses occupational therapists’ (OTs) shared decision- making (SDM) behaviors with individuals having persistent pain and explored factors influencing SDM behaviors. | Cross-sectional study | Third party observer of consultations using OPTION and DCS. | 15 HCP (occupational therapists)  37 patients.. |
| **19** | Dima, 2013, England | British Journal of General Practice | Identifying patients' beliefs about treatments for chronic low back pain in primary care: a focus group study | To explore patient preferences and to identify patients’ beliefs about LPB treatments. | Qualitative | Focus groups interviews | 75 patients with LBP. |
| **110** | Eilayyan, 2018, Canada | BMC Musculoskeletal Disorders | Promoting the use of self-management in novice chiropractors treating individuals with spine pain: the design of a theory-based knowledge translation intervention | To 1) estimate the organizational readiness for change toward using SMS at the Canadian Memorial Chiropractic College (CMCC), Toronto, Ontario from the perspective of directors and deans, 2) estimate the attitudes and self-reported behaviours towards using evidence-based practice (EBP), and beliefs about pain management among supervisory clinicians and chiropractic interns, 3) identify potential barriers and enablers to using SMS, and 4) design a theory-based tailored Knowledge Translation (KT) intervention to increase the use of SMS. | Mixed method | Questionnaires and focus group interviews | 141 HCP’s (decision makers, clinicians and chiropractic interns). |
| **111** | Eilayyan, 2019, Canada | Chiropractic and Manual Therapies | Promoting the use of self-management in patients with spine pain managed by chiropractors and chiropractic interns: barriers and design of a theory-based knowledge translation intervention | To: 1) assess participation in self-care (i.e. activation) among patients with spine pain, 2) identify patients’ barriers and enablers to using SMS, and 3) map behaviour change techniques (BCTs) to key barriers to inform the design of a knowledge translation (KT) intervention aimed to increase the use of SMS. | Mixed method | Questionnaires and semi-structured individual interviews | 223 patients with spine pain. |
| **112** | Fenety, 2009, Canada | Manual Therapy | Informed consent practices of physiotherapists in the treatment of low back pain | To explore and describe physiotherapists’ informed consent practices in the treatment of clients with low back pain | Qualitative | Focus groups | 44 HCP’s (physiotherapists). |
| **113** | Fraenkel, 2007, USA | The Journal of Rheumatology | Improving informed decision-making for patients with knee pain | To test the efficacy of a computer tool to improve informed decision-making for patients with knee pain in an outpatient primary care clinic setting. | Randomized controlled trial | Questionnaires | 87 patients with knee pain. |
| **114** | Fu, 2018, England | Qualitative Health Research | The Management of Chronic Back Pain in Primary Care Settings: Exploring Perceived Facilitators and Barriers to the Development of Patient–Professional Partnerships | To understand patient–professional partnership and its related factors that may facilitate or hinder patients’ self-management through exploration of patients’ experiences and perceptions for pain management in primary care settings. Findings reported here are based on secondary analysis of the qualitative phase of a larger mixed-methods study that evaluated the impact of patient–professional partnerships on self-management of chronic pain. | Qualitative | In-depth, semi structed interviews | 26 patients. |
| **115** | Hochlehnert, 2006,  Germany | Patient Education and Counselling | A computer-based information-tool for chronic pain patients. Computerized information to support the process of shared decision-making. | Assessment of the use of a computerized information-tool in the context of a shared decision-making process with chronic pain patients. | Mixed methods prospective randomized controlled trial | Questionaries (DCS and SWD)  Semi-structures interviews | 75 patients with fibromyalgia. |
| **216** | Ibrahim, 2017, USA | JAMA Surgery | Effect of a Decision Aid on Access to Total Knee Replacement for Black Patients With Osteoarthritis of the Knee: A Randomized Clinical Trial | To assess whether a decision aid improves access to total knee replacement (TKR) surgery for black patients with OA of the knee. | Single-blinded randomized clinical trial | Receipts of TKR within 12 months and questionnaires | 336 patients with knee pain. |
| **217** | Jansen-Kosterink, 2021, Netherlands | BMC Medical Informatics and Decision Making | Clinician acceptance of complex clinical decision support systems for treatment allocation of patients with chronic low back pain | To identify the factors (and their importance) that hinder or alleviate the acceptance of clinicians toward the use of a complex CDSS for treat- ment allocation of patients with chronic low back pain. | Mixed methods (quantitatively driven study) | Questionnaire | 89 HCP’s (general practitioners, primary care physical therapist, clinicians at rehabilitation centres). |
| **218** | Jones, 2022, United States | Journal of Family Medicine and Primary Care | Development of a personalized shared decision-making tool for knee osteoarthritis and user-testing with African American and Latina women | To analyze the patient experience with a personalized knee osteoarthritis SDM tool in African American and Latina women | Parallel-designed, double blinded randomized controlled study | Questionaries and interviews | 104 patients with knee arthritis. |
| **219** | Kravitz, 2018, USA | JAMA Internal Medicine | Effect of Mobile Device-Supported Single-Patient Multi-crossover Trials on Treatment of Chronic Musculoskeletal Pain: A Randomized Clinical Trial | To determine whether patients randomized to participate in an n-of-1 trial supported by a mobile health (mHealth) app would experience less pain and improved global health, adherence, satisfaction, and shared decision making compared with patients assigned to usual care. | Randomized clinical trial | Questionnaires | 215 patients with chronic musculoskeletal pain. |
| **220** | Laerum, 2006, Norway | Journal of Rehabilitation Medicine | What is "the good back-consultation"? A combined qualitative and quantitative study of chronic low back pain patients' interaction with and perceptions of consultations with specialists | To identify and describe the core elements (and style) of what patients with chronic LBP perceive as good clinical communication and interaction with their doctor (specialist) in a consultation, and to explore whether there were any special characteristics of consultations per- formed by doctors using consultation models with previously documented effect (return to work). | Qualitative (with some quantitative data) | Observing of consultations  Interviews | 35 patients with chronic low back pain.  14 HCP’s (specialists in neurology, rehabilitation medicine, orthopaedics, neurosurgery and rheumatology). |
| **21** | Li, 2025, China | Patient Preferences and Adherence | “Doctor-Led, Patient-Centered”: A Mixed-Method Research Comparing Patients’ and Doctors’ Treatment Outcome Choices for Chronic Low Back Pain | This study aims to analyze the alignment and differences in doctors’ and patients’ perceptions of treatment outcomes and explore the implications of these differences | Explanatory sequential mixed-methods design | Questionnaire and focus groups | Questionnaire: 30 patients with cLBP and 26 doctors.  Focus group:  8 patients with cLBP and 8 doctors. |
| **22** | Licciardone, 2024, USA | The Journal of Pain | Impact of Shared Decision-Making on Opioid Prescribing Among Patients With Chronic Pain: A Retrospective Cohort Study | This study measured whether SDM affects opioid prescribing frequency for chronic low back pain | Retrospective Cohort Study | Questionnaires | 1478 patients with chronic pain. |
| **23** | Leininger, 2025, USA | Chiropractic & Manual Therapies | Supported biopsychosocial self-management for back-related leg pain: a randomized feasibility study integrating a whole person perspective | The objective was to assess feasibility using pre-specified targets for recruitment and enrollment, intervention delivery and data collection. | A randomized feasibility study | Surveys and interviews | 42 patients with back-related leg pain. |
| **223** | Merolli, 2019, Australia | Studies in Health Technology and Informatics | User-Centered Value Specifications for Technologies Supporting Chronic Low-Back Pain Management | To investigate what will be required for participatory health enabling technologies for them to have a significant effect on cLBP management. | Qualitative | Semi-structered interviews | 10 HCP’s (clinical health professionals with expertise in managing cLBP).  10 patients living with cLBP. |
| **222** | Parsons, 2012, England | Family Practice | Will shared decision making between patients with chronic musculoskeletal pain and physiotherapists, osteopaths and chiropractors improve patient care? | To explore patients and chiropractors, osteopaths and physiotherapist’s beliefs about CMP and its treatment and how these beliefs influences care seeking and ultimately the proces of care. | Qualitative | In-depth interviews and focus groups | 13 patients with CMP.  19 primary care health professionals (osteopaths, chiropractors, physiotherapists). |
| **323** | Patel, 2014, England | BMC Muscoloskeletal Disorders | Primum non nocere: shared informed decision making in low back pain -- a pilot cluster randomised trial | The purpose of this study was to pilot a decision support package to help people choose between low back pain treatments | Cluster randomised trial | Questionaries | 19 HCP’s (physiotherapists).  148 patients. |
| **224** | Rashidian, 2013, Iran | BMC Research notes | The perspective of iranian physicians and patients towards patient decision aids: a qualitative study. | To describe physicians’ and patients’ viewpoints on the barriers and limitations of using patient decision aids in Iran, their proposed solutions, and, the benefits of using these tools | Qualitative | Interviews | 14 physicians.  8 patients. |
| **325** | Riffin, 2015, USA | Journals of Gerontology: Social Sciences | Decision Support Preferences Among Hispanic and Non-Hispanic White Older Adults With Chronic Musculoskeletal Pain | To obtain qualitative data on whether and why culturally diverse CMP patients turn to relatives, friends, and health care providers when making decisions about their pain care. Specifically, we explore (a) the types of support (emotional, instrumental, and informational) patients desire and receive from formal and informal network ties and (b) patients’ perceptions of and satisfaction with the support they receive. Ethnic differences and decision characteristics are considered throughout. | Qualitative | Semi-structured interviews | 63 participants (Hispanic and non-Hispanic older white adults living with CMP). |
| **326** | Sanders, 2018, Netherlands | BMC Family Practice | The effectiveness of shared decision-making followed by positive reinforcement on physical disability in the long-term follow-up of patients with nonspecific low back pain in primary care: a clustered randomised controlled trial | To assess the effectiveness of shared decision-making followed by a positive reinforcement of the chosen therapy (SDM&PR) on patient-related clinical outcomes in patients with non-chronic low back pain in general practice. | Cluster-randomised controlled trial | Questionnaires | 68 HCP’s (GP’s).  226 patients with non-chronic non-specific low back pain. |
| **327** | Saunders, 2020, England | BMC Family Practice | Stratified primary care versus non-stratified care for musculoskeletal pain: qualitative findings from the STarT MSK feasibility and pilot cluster randomized controlled trial | To explore the feasibility of delivering stratified care ahead of the main trial. | Qualitative | Stimulated -recall interviews  Video/audio-recording of consultations with stratified care. | 10 patients.  10 GP’s. |
| **328** | Stenner, 2016, England | Physiotherapy | Exercise prescription for patients with non-specific chronic low back pain: a qualitative exploration of decision making in physiotherapy practice | To explore how shared decision making and patient partnership are addressed by physiotherapists in the process of exercise prescription for patients with NSCLBP. | Qualitative | Observation field notes and semi-structures interviews | 8 physiotherapists. |
