## Appendix 4 for "Implementation of Shared Decision-Making in the Management of Chronic Musculoskeletal Pain: *a scoping review*"

1.1. **Patient-level factors**
     1.1.1. ***Cultural differences***

This theme was only represented by patients and reported as both a barrier and a facilitator. Facilitators [40,66] were related to culturally competent providers who offered emotionally tailored decisions and decision aids tailored to provide patient education [40]. Furthermore, micro-cultural differences were reported as both a barrier and facilitator to SDM, as cultural differences influenced trust in one’s doctor and participation in decision-making [11].

1.1.2. ***Decision support***

This theme was only represented by patients and generated three subthemes: “turning to relatives”*,* “turning to peers”, and “turning to friends/network”*.* The subtheme “turning to relatives” was reported as both a barrier and facilitator. Barriers were related to pressure from relatives, or the patient selecting treatment options not aligning with relatives’ beliefs, leading to dissatisfaction with the decision [66]. A facilitator was emotional reassurance by relatives [66]. Factors considered both a barrier and facilitator were the need for relatives to reassure the patient whether the right decision was being made, especially in decisions with the potential to compromise the individual’s functional ability [66]. The subtheme “turning to peers” was reported as both a barrier and a facilitator [66]. A facilitator was related to talking to others with similar pain guiding treatment choices. Furthermore, it was considered both a barrier and facilitator that some patients only wanted advice from others with similar pain to guide their decisions. The subtheme “turning to friends/network” was considered both a facilitator and a barrier [66]. Facilitators were related to reaching out to friends with medical training or informal guidance. Considered both a barrier and facilitator were when patients used formal network channels regarding high-stake treatments.

1.1.3. ***Individualized care***

This theme was represented by patients and HCPs and only had one sub-theme, “Concern of adverse events”. Facilitators to SDM [17,20,32] were related to support adjusted to the different stages of treatment [32], management and goal setting congruent with lifestyle, personal needs, and life situation, as well as addressing other issues than the pain only [10]. Receiving a diagnosis was reported as both a barrier and facilitator by patients [10], since receiving a diagnosis could be perceived as a relief to some, but to others as insufficient, labelling, and leading to standardized care instead of individualized care [10]. Patient barriers [17,32,47] were related to a lack of individualized care [32], especially in settings with group rehabilitation [17]. It was considered a barrier when HCPs did not focus on how the pain affected psychosocial factors and vice versa during consultations (e.g., daily life situations, including job, family, quality of life, etc.) [47]. Both patients and HCPs expressed concerns about adverse events for different interventions related to the individual [52]. Importantly, these concerns often differed between patients and HCPs, depending on the treatment acting as a barrier for SDM [52].

1.1.4. ***Knowledge about disease, condition, options, and outcomes***

This theme was primarily represented by patients. Six subthemes emerged: “Patient education”, “explanation of pain”, “lack of diagnostic certainty”, “belief of pain origin”, “providing information about options”, “expectations of outcome”, and “navigating in uncertainty relating to inadequate treatment provision”. Both patients [18,23,75] and HCPs [56] considered the sub-theme “patient education” a facilitator to SDM, fostering an improved understanding of their own condition. Patients considered educational materials and resources, anatomical and pathophysiological explanations in layman’s terms during consultation and examination facilitators to SDM [10,17,18,20,23,47,56,75]. HCPs also considered educational materials a facilitator to SDM [56], as well as patient education, increased cooperation, and engagement in treatment decisions [65]. HCPs reported barriers to SDM when they felt their explanations had not gone far enough. Furthermore, patients considered it a barrier [75] when they were too knowledgeable to obtain any new knowledge in the SDM encounter. Patients reported it both a barrier and a facilitator when engaging in SDM could be dependent on whether they believed a treatment was credible and made sense [7] or if their belief of pain origin aligned with the HCP’s (e.g., strong biomedical views vs acceptance of psychological explanations) [20,61]. “Explanation of pain” emerged as a subtheme reported primarily by patients. Patients only reported this subtheme as a barrier to SDM [17,47,61], with influencing factors being a lack of explanation of pain causation and inconsistent explanations of their pain, leading to frustration and adaptation of different beliefs of pain causation as they saw different HCPs. HCPs also reported inconsistent explanations of pain as a barrier to SDM [54]. In continuation, “lack of diagnostic certainty” was an emerging subtheme reported by both patients and HCPs. Patients reported barriers to be when there was no specific cause of pain, when they felt under pressure to convince HCPs of their pain, and when no new advice was given during consultations [47,61]. However, receiving a diagnosis was also considered a facilitator when HCPs asked what previous advice was given [61]. Adversely, HCPs considered it a facilitator when patients trusted their clinical judgment even when the patient’s pain was unspecified [47,61]. The subtheme “belief of pain origin” was primarily represented by patients and was considered both a barrier and facilitator for SDM. Patients reported it as both a barrier and facilitator whether the HCP’s and their beliefs of pain origin aligned [61]. Barriers were related to the HCP’s and patient’s beliefs about the causation of pain, when non-congruent, which could lead to the patient feeling the HCP delegitimised their pain, whereas congruent pain beliefs and explanations facilitated SDM. Furthermore, in time, patients would accept or reject pain explanations, whether they were congruent with their own pain beliefs [61]. In continuation hereof, with different beliefs of pain origin, patients reported barriers such as HCPs believing the patients are “faking it”, age-related stigma, and patients being concerned that their clinician believed they are hypochondriacs [10,20]. HCPs considered explaining psychological aspects of pain a barrier to SDM [61] when there was a dissonance in the patients’ and HCPs’ beliefs of psychosocial aspects of care regarding the pain, which could lead to an impaired patient-HCP relationship. The subtheme “providing information about options” was represented primarily by patients. Patient-related facilitators [20,47,66] were related to empathetic interactions [66], a systematic active approach evaluating initial effectiveness of individualized care [20], and sufficient information for them to make a decision [47]. Observers reported clear information from the HCP when serious diseases could be ruled out as a facilitator to SDM [47]. The delivery of treatment options that were considered credible and delivered by the right practitioner was considered both a barrier and facilitator to SDM by patients [20]. Barriers reported by patients were related to lack of explanation of what options were available and not knowing the cause of the pain [17]. An HCP barrier was insecurity about whether explanations had gone far enough [75]. The subtheme “expectations of outcome” was primarily reported by patients. Factors considered both facilitators and barriers by patients [23,47,61] were patients' self-perception and age-related expectation of health [47], belief of pain origin (biomedical vs psychological) [61], and emotions concerning the use of SDM [23]. Observers reported it a barrier when HCPs explored patients’ expectations and preferences without taking it into account, but also when there was a mismatch between the doctors’ and patients’ expectations [47]. HCPs reported it is a barrier to managing unrealistic patient expectations [61]. The subtheme “navigating in feeling giving inadequate treatment provision” was reported as a barrier by HCPs [61,75]. Influencing factors were the pressure of “wanting to give the patient something” and the pressure to provide a diagnosis without the ability to provide different treatment offers than previously suggested [61].

1.1.5. ***Patient characteristics***

This theme was represented by both HCPs and patients and generated one subtheme: “managing/explaining psychological aspects of pain”. Generally, patient characteristics were reported as a barrier to SDM by both patients and HCPs. Patient barriers [10] were related to patient resources, specifically not having resources to lead the course of management or when treatment options became unavailable due to economy. HCP barriers [7,22] of patient’s characteristics were related to patients with psychological overlay, lack of compliance, lack of adherence to SDM, lack of motivation, level of education, resources, not accepting the condition, not trusting the clinician, language- or cultural barriers, and lastly not wanting to use SDM. The subtheme “managing/explaining the psychological aspects of pain” was represented by patients and HCPs and considered both a barrier and a facilitator. Patients considered discussing the psychological aspect of pain both a facilitator [61,69] and a barrier [10,47,61]. Patient barriers [10,61] were related to feeling that psychological explanations delegitimized their pain, and not buying into psychological explanations of their pain. In continuation hereof, the patients' extent of biomedical view was also considered a barrier since patients with strong biomedical causal explanations were not willing to accept psychological models as the cause of pain [61]. However, patients also considered it a barrier when HCPs did not discuss the possible correlation of psychosocial issues, not acknowledging the pain, and its influence on daily life [47].

1.1.6. ***Time***

This theme was represented by both patients and HCPs and generated three subthemes: “workflow”, “time duration with MSK pain”, and “time pressure”. The subtheme “workflow” was only represented by patients [10,17,23,32]. Barriers were short appointment times [10,32,53] and long waiting times [32]. Longer consultation time was considered a facilitator [18]. The subtheme “time duration with MSK pain” was represented by both patients and HCPs. Patients considered it both a barrier and facilitator [61] to incorporate age-related perception of their own health and pain acceptance (i.e., their perception of how their health should be at a certain age). Furthermore, incorporating physical and psychological explanations dependent on pain duration and length of therapeutic relationship was considered both a barrier and facilitator by patients and HCPs [61]. The last subtheme, “time pressure”, was also represented by HCPs and patients and was only considered a barrier to SDM [10,22,29,32,41,65]. HCP-related barriers were influenced by limited time since SDM was thought to be more time-consuming. Patients reported that time pressure was a constraint in building a partnership with the HCP.

1.1.7. ***Trust***

This theme was reported by both patients and HCPs and generated two subthemes: “stigma” and “pain legitimation”. In general, trust was reported by both patients and HCPs as both barriers and facilitators. Patient barriers to SDM were [32] losing trust in their HCPs who offered previously suggested treatment management, which was not effective. Patient facilitators [10,11,17] were emotional support [66], taking time to discuss management [66], empathetic interactions [17,66], and being honest and open to help the HCP identify relevant treatment options within a limited time [10]. HCP facilitators were building rapport [29], and barriers were patients not trusting their HCP [22]. The last subtheme, “pain legitimation” (i.e., acknowledging the patient's experience of pain), was represented by both patients and HCPs. Both HCPs and patients reported pain legitimation as a facilitator of SDM [61]. Patient facilitators were knowledgeable, respectful, and trustworthy HCPs who believed and acknowledged the individual’s experience of pain [20,32]. Patient barriers to SDM [47] were the pressure of convincing the doctor of their pain, feeling disbelieved, and being told to live with the pain. HCP facilitators were related to the HCP expressing empathy and believing that the pain was real [47].

1.2. **Clinician-level factors**
     1.2.1. ***Attitude and perspectives towards shared decision-making***

This theme was primarily represented by HCPs [7,10,22,23,29,41,54,61,69,75]. Five subthemes were generated: “quality of SDM”, “perceived usefulness”, ”threat to autonomy”, “lack of self-efficacy”, and “observability of SDM intervention”. The subtheme “quality of SDM” was considered both a barrier [41] and a facilitator [7]. The subtheme “perceived usefulness” was primarily considered a facilitator [22,23,54]. Facilitators were believing in and feeling optimistic that SDM would lead to improved patient outcomes. The intention to use SDM was also reported as both a facilitator and barrier [41], where the barrier was related to the belief of increased service risks (i.e., SDM was considered inferior to the existing situation). However, a barrier to implementing and using SDM in clinical practice was disagreement with the principles of SDM and choosing paternalistic care approaches [75]. The subthemes “threat to autonomy” and “lack of self-efficacy” were considered barriers to SDM. Threat to autonomy was reported as worries of undermining professional autonomy using SDM [29,41,75]. Lack of self-efficacy was influenced by not feeling competent to perform SDM, not providing sufficient explanations, lack of skills, and anxiety using SDM on patients with psychological overlay [22,61,75]. Facilitators related to the subtheme “observability of SDM intervention” were when HCPs were able to observe patients benefit from SDM interventions [22], and barriers were lack of evaluation of SDM interventions, leading HCPs to apply it based on their own preferences [65].

1.2.2. ***Characteristics of HCP***

This theme was only reported by patients and did not generate any subthemes. Patients considered it a barrier when HCPs were abrupt during the encounter [17]. Additionally, patients considered the length of the relationship with the HCP as both a barrier and a facilitator [61]. Patient-related facilitators were related to empathetic behaviour, showing emotional support, demonstrating professional and respectful behaviour, and acknowledging and believing the patient’s pain experience [17,20,32,66].

1.2.3. ***Communication***

This theme was represented by both HCPs and patients and generated four subthemes: “guiding the patient through the consultation”, “terminology used by the clinicians”, “non-verbal communication”, and “language barriers”. In general, both patients and HCPs reported communication as both a facilitator and a barrier, dependent on communication techniques. Patient-related facilitators were influenced by well-explained treatments and the quality of communication [32,53]. HCP-related facilitators were influenced by SDM training to ease interaction and patients confirming they agree to the explained treatment options [29]. Communication barriers for HCPs were considered lack of communication skills [22,53]. The subtheme “guiding the patient through the consultation” was represented by patients, HCPs, and observers and was considered both a facilitator and a barrier. Patient facilitators [17,47] were communication with good explanations, enabling patients to draw conclusions on their own [17,47] and explanations during examination of what was done and why [47]. Observer facilitators were communicating positive findings/positive feedback and effective reassurance, ruling out serious disease at an early stage [47]. Lack of guidance during the consultation was reported by patients as a barrier, particularly when insufficient explanations led to uncertainty about the origin of their pain and the available treatment options [17,47,69]. HCP barriers were not guiding to prevent confusion among the patients [29] and lack of HCP training within SDM, especially in complex conditions [53]. The subtheme “terminology used by clinicians” was represented by both patients and observers. Patient facilitators [17,32,47] occurred when HCPs used layman terms [17,32] and provided understandable explanations consistent with the patient’s health literacy level [47]. Observer facilitators were simple explanations and metaphors when explaining health topics [47]. Patient barriers were use of advanced medical terms and diagrams [17,47]. The subtheme “non-verbal communication” was only reported by HCPs as a barrier, where reliance on non-verbal cues, such as reduced eye contact or posture, made it difficult to engage in SDM [29]. Instead of fostering open discussion, these cues were sometimes interpreted by HCPs as indicators of patient consent or discomfort, leading to assumptions about treatment preferences without fully involving the patient in the decision-making process [29]. The subtheme “language barrier” was reported by patients and HCPs as both a barrier and a facilitator. Patient barriers were a lack of in-depth treatment discussion when not in the patient’s first language and lack of high-quality interpreter services [66]. HCP considered language barrier leading to restriction in SDM use [22]. One patient facilitator was [66] the linguistic compatibility of the provider and the patient.

1.2.4. ***Knowledge of the application and use of shared decision-making***

This theme was primarily represented by HCPs. Four subthemes were generated: “familiarity with SDM”, “assumed efficacy vs informed choices”, “timing of shared decision making”, and “clinical situation”. The subtheme “familiarity with SDM” was only represented by HCPs [22,23]. Barriers were related to the lack of comprehensive SDM courses to obtain familiarity with SDM [22] and facilitators were related to pre-existing knowledge of SDM [22,23]. The “subtheme assumed efficacy vs informed choices” was reported only by HCPs and only as a barrier [29,41,53,75]. Barriers to SDM were HCPs choosing management plans based on their personal preferences rather than informed patient choices [29,41,75] and the use of an adopted patient-centred approach to obtain compliance with their expert recommendations [75]. The subtheme “timing of SDM” was reported as both a facilitator and barrier, where the best timing of SDM was perceived differently between patients and HCPs [1,29]. Facilitators to SDM were related to the degree of the treatment decision-complexity, whereby high-risk treatments occasionally promoted greater SDM, while low-risk decisions hampered SDM and favoured informed consent. On the contrary, barriers were related to HCPs relying on the patients to refuse treatment rather than obtaining consent. Another barrier was related to HCPs using intuition to select which patients SDM should be applied to [23]. The subtheme “clinical situation” was only represented by HCPs. Barriers were related to decision aids being too difficult to use [41,75] and not finding SDM suitable for the individual clinical situation [29,69]. However, using decision aids was also considered a facilitator to SDM as decision aids could be used as a checklist to confirm all suitable management options were considered and nothing was overlooked [69].

1.2.5. ***Skills***

This theme was represented by both patients and HCPs. Two subthemes were generated: “SDM training” and “therapeutic alliance”. Formal training in SDM was considered a facilitator by both HCPs [6,7,18] and patients [6,7,10]. However, a lack of formal SDM training was reported as a barrier to performing SDM by HCPs [10,22]. Furthermore, patients also identified it as a barrier when SDM training of HCPs did not lead to satisfactory implementation, as patients did not report a significant improvement in treatment satisfaction following SDM [10,62]. The subtheme, obtaining a good therapeutic alliance, was primarily considered a facilitator to SDM by patients [6,7,10,20,32,51] and HCPs [6,7,29,32,68]. Factors contributing to a good therapeutic alliance reported by patients included feeling understood, the degree of involvement in SDM, having a sustained relationship with their HCP, and the competence of the HCP. For HCPs, factors such as positive feelings towards the patients, SDM training, and trust contributed to an enhanced therapeutic partnership, which facilitated SDM. Conversely, the lack of obtaining a therapeutic alliance was considered a barrier to SDM by both patients [10,61] and HCPs [29]. HCPs reported influencing factors as not providing sufficient information during the consultation and under confidence in delivering psychological aspects of pain to patients.

1.3. **Interactional dynamics**
     1.3.1. ***Decision aid***

This theme was related to multiple different decision aids and was represented by both patients and HCPs. The theme generated four subthemes: “facilitating preparedness to participate in SDM using decision aids”, “decision aids features and user-experience”, “checklist diagnosing”, and “wording of decision aid”. In general, decision aids were primarily considered a facilitator to SDM by both patients and HCPs. HCP facilitators [69] were related to adopting a more functional approach, meaning the HCPs were not expected to solve the pain manner, being able to provide several treatment options, and facilitate the discussion of psychological concerns. HCP barriers were related to decision aids hampering personal contact with the patient [47] and being time-consuming [65]. Patient facilitators [38,40,43,46,69] were related to a more thorough and effective consultation with the HCP [69], facilitating patient education and knowledge on treatment options [38,40,43], improving the partnership and trust with HCPs [46], increased confidence in patient management [46] and perceived usefulness [38], and prevention of unnecessary referrals [65]. Furthermore, patients reported age in relation to decision aids as both a barrier and facilitator to SDM, since older patients needed further support to navigate a decision aid [38]. The subtheme “facilitating preparedness to participate in SDM using decision aids” was primarily represented by patients and only considered a facilitator [14,30]. Patient-perceived facilitators included decision aids that reduced decisional conflict, enhanced self-efficacy, and helped prepare patients to actively participate in decision-making. HCPs reported patients becoming more psychologically ready for decision making [65]. The subtheme “decision aid features and user experience” was reported by both patients and HCPs and was considered a facilitator. Patient facilitators [43,56] were related to the design/features of a decision aid, which were related to tracking metrics, alerts reinforcing positive behaviour, improved user experience, and decision aids that were non-complicated to use. Both patients and HCPs [56] reported robust communication features (i.e., SMS, email, secure messaging features) and educational materials (i.e., videos) as facilitators. The subtheme “checklist diagnosing” was represented by both HCPs and patients. HCP-related facilitators were related to confirming all suitable management options were considered and supporting and sharpening diagnostic thinking [41,69]. However, a barrier for HCPs was that the decision aid was too generic, not allowing for different patient characteristics [41]. A patient-related facilitator was trusting the HCP and only asking questions of relevance [69]. The last subtheme, “wording of decision aids” was represented by both HCPs and patients and was only considered a barrier. A patient barrier was closed questions restricting open discussion with the HCP [69]. HCP barriers were related to awkward wording of questions, not fitting into the natural flow, or wording of questions leading to misunderstandings or unusual answers [69].

1.3.2. ***Decision dynamics: Navigating patient-HCP roles, values, and power***

This theme was represented by both HCPs and patients and generated five subthemes: “decision characteristics”, “sharing responsibility”, “patient wants paternalistic decision-making”, “asking patients about their preferred role in decision-making”, and “asking patients about values/clarifying values”. The subtheme “decision characteristics” was represented only by patients and was considered both a barrier and a facilitator. Barriers to SDM were related to high-stakes decisions where patients turned to relatives or others; however, turning to others at high-stakes decisions was also reported as a facilitator to SDM [66]. The subtheme “sharing responsibility” was represented by both HCPs and patients. Patient-related facilitators [23,32] were related to involvement in decision-making leading to motivation [23], and that involvement led to increased mutual understanding and trust [32]. Patient-related barriers were being disrespected or disbelieved by their HCP [20]. HCPs reported patients’ willingness to take responsibility for their condition as both a barrier and facilitator [61], since it also impacted their care-seeking behaviour, however, decision aids were shown to facilitate patients taking responsibility in their treatment decisions. HCP-related facilitators were related to patient empowerment [41,75]. The subtheme “patients want paternalistic decision-making” was considered a barrier by patients and HCPs. HCP-related barriers were patients actively not wanting to participate in decision-making [7,22], not wanting SDM due to lack of familiarity with their own rights [65], or shifting from a traditional provider-driven approach [51]. Patient-related barriers were related to believing the HCP as the only expert, and that the HCP, therefore, should decide the management course [10,17,66,69]. The subtheme “asking patients about their preferred role in decision-making” was reported by patients and HCPs. Patient facilitators were related to involvement in decision making, which led to motivation [23] and increased trust and confidence in their HCP [65]. Patient barriers were not being asked about their preferred role and involvement in decision-making [61]. HCPs reported it a barrier since asking the patient could lead to a misunderstanding that the physician did not have enough knowledge or experience [65]. The subtheme “asking patients about values/clarifying values” was reported by both HCPs and patients [61]. Both HCPs and patients reported continuous revisiting of the patient’s beliefs during treatment as a facilitator [61], as well as taking the patient's autonomy into consideration [65]. HCP facilitators were related to considering patients’ values and perspectives, which led to engaging and motivating patients [75]. HCP-related barriers were related to patients not being asked about values and just offering default treatment options [75].

1.4. **System-level factors**
     1.4.1. ***Cost-beneficial***

The cost-beneficial aspect of SDM was a barrier reported by both HCPs [22,41] and patients [10,41,62]. HCP perceived barriers included lack of effectiveness and added value, risk of losing patients wanting passive care, and increased costs and time. Patient-related barriers were increased time and/or costs. However, HCPs also reported decision aids as a facilitating SDM intervention due to their reduction of workload and time [41]. Both patients and HCPs reported that decision aids were believed to reduce costs by improving decision accuracy in the first consultation and decreasing incorrect decisions, thereby reducing costs [65].

1.4.2. ***Healthcare system organization***

This theme was represented by both patients and HCPs and generated six subthemes: “Characteristics of healthcare setting”, “healthcare process expectancy”, “lack of resources”, “lack of access”, “organisational constraints”, and “multidisciplinary care”. The “characteristics of the healthcare setting” were considered both barriers and facilitators by patients and HCPs. Patient-related facilitators [17,23,69] were thorough assessments [17], efficient consultations [69], cooperation by HCPs and a homely environment [23]. Patient barriers [10,17,20] were negotiation of referrals and tests [20], cost of care [10], organization of appointments [17] and busy unit/rushed environment [10,17]. Factors considered both barriers and facilitators by patients were service and system-related factors and healthcare setting (e.g., GP vs pain clinic) [32]. For HCPs the clinic characteristics (e.g., equipment, sufficient space, etc.) were considered both a facilitator and a barrier [22]. HCP barriers were economic benefits to discard choosing SDM [65]. The subtheme “healthcare process expectancy” was considered a barrier by HCPs and patients, believing that SDM would not lead to an improved healthcare process [69]. HCP-related barriers were believing that using SDM would confuse the patient, choosing their own preferred management rather than the patient's, risk of losing patients, and not referring patients due to lack of local specialised healthcare services [22,75]. The ladder was also considered a barrier for patients [69]. The subthemes “lack of resources”, “lack of access”, and “organizational constraints” were all considered barriers by HCPs and patients. In relation to “lack of resources”, HCP barriers [22] were financial constraints, lack of specialists per capita, and increased workloads. Both HCPs and patients reported resource constraints as a barrier [32]. Barriers regarding the subtheme “lack of access” were reported by HCPs and patients. Both reported lack of access to services [69] as a barrier. HCP barriers included lack of access to formal SDM training [10,22]. The subtheme “organizational constraints” was reported by HCPs as both a barrier and a facilitator and was related to the influence of others (e.g., supervisors, colleagues) to use SDM [22,23], since superiors’ attitude towards SDM either facilitated or restricted the use of SDM. The subtheme “multidisciplinary care” was only represented by patients and was considered both a barrier and facilitator to SDM. Facilitators were multidisciplinary care in general and having goals and solutions discussed with HCPs with clinical expertise in the field [10,32]. However, a barrier [32] was not being able to develop a proper partnership with each HCP within the multidisciplinary team.
